# Descriptive Epidemiological Assessment of the Relationship between the Global Burden of Influenza from 2017-2019 and COVID-19

**DOI:** 10.1101/2020.06.18.20134346

**Authors:** Stefan David Baral, Katherine B. Rucinski, Jean Olivier Twahirwa Rwema, Amrita Rao, Neia Prata Menezes, Daouda Diouf, Adeeba Kamarulzaman, Nancy Phaswana-Mafuya, Sharmistha Mishra

## Abstract

**Background:** SARS-CoV-2 and influenza are lipid-enveloped viruses with differential morbidity and mortality but shared modes of transmission.

**Objective:** With a descriptive epidemiological framing, we assessed whether recent historical patterns of regional influenza burden are reflected in the observed heterogeneity in COVID-19 cases across regions of the world.

**Methods:** Weekly surveillance data reported by the World Health Organization from January 2017– December 2019 for influenza and through October 31, 2020 for COVID-19 were used to assess seasonal and temporal trends for influenza and COVID-19 cases across the seven World Bank regions.

**Results:** In regions with more pronounced influenza seasonality, COVID-19 epidemics have largely followed trends similar to those seen for influenza from 2017-2019. COVID-19 epidemics in countries across Europe and Central Asia and North America, have been marked by a first peak during the spring, followed by significant reductions in COVID-19 cases in the summer months, and a second wave in the fall. In Latin America and Caribbean, COVID-19 epidemics in several countries peaked in the summer, corresponding to months with highest influenza activity in the region. Countries from regions with less pronounced influenza activity including South Asia and Sub-Saharan Africa, showed more heterogeneity in COVID-19 epidemics seen to date. However, similarities in COVID-19 and influenza trends were evident within select countries irrespective of region.

**Conclusion:** Ecological consistency in COVID-19 trends seen to date with influenza trends suggest the potential for shared individual, structural, and environmental determinants of transmission. Using a descriptive epidemiological framework to assess shared regional trends for rapidly emerging respiratory pathogens with better-studied respiratory infections may provide further insights into the differential impacts of non-pharmacologic interventions and intersections with environmental conditions. Ultimately, forecasting trends and informing interventions for novel respiratory pathogens like COVID-19 should leverage epidemiologic patterns in the relative burden of past respiratory pathogens as prior information.

## Introduction

The COVID-19 pandemic has affected each region of the world differently. As of December 2020, Europe and the Americas regions have registered the highest numbers of COVID-19 cases and deaths while South-East Asia, Sub-Saharan Africa, and the Western Pacific have experienced comparatively milder epidemics [1]. Heterogeneity in COVID-19 burden has also been observed within cities, across cities, and across regions within a country [2, 3]. This heterogeneity likely stems from differences in underlying population structure with respect to age distribution, population and housing density, access to health care, burden of comorbidities, socio-economic and structural barriers to engagement in health, environmental factors, and the breadth and intensity of public health interventions. Furthermore, there remains active investigation into the heterogeneity of effects of both COVID-19 and the associated response on potential syndemics including infectious diseases such as HIV and tuberculosis and non-communicable conditions including mental health, reproductive health, and malnutrition [4].

Similar to SARS-CoV-2, the virus that causes COVID-19 disease, influenza is a lipid-enveloped respiratory virus but with a more established evidence base for both effective non-pharmacologic intervention strategies and transmission dynamics [5-7]. Importantly, influenza is associated with a shorter incubation period and with lower mortality than the currently estimated SARS-CoV-2 infection case fatality rate [8-10]. The relative risks of indoor vs. outdoor transmission, risks for contact, droplet-based, and aerosolization-mediated transmission are better established for influenza [11-13]. Influenza seasonality and spatial distribution are not fully understood but likely reflect individual, population, and environmental determinants [14]. Moreover, the annual burden and associated mortality secondary to pandemic and non-pandemic seasonal influenza strains vary significantly. The 2009 H1N1 pandemic was estimated to cause over 60 million infections in the United States, and resulted in higher mortality among youth but fewer deaths overall as older people were more likely to be immune [15]. In 2017, there were an estimated 45 million cases of mostly H3N2 and Influenza B in the United States associated with higher mortality particularly among older adults given limited pre-existing immunity and limited vaccine effectiveness [16]. Environmental factors have been shown to explain, in part, the differences in influenza seasonality between temperate regions where cases peak during winter months and tropical regions which may have multiple peaks or intermittent influenza activity [17, 18]. However, even within specific regions, some countries show influenza transmission patterns that appear distinct from regional trends [19, 20].

There have been early reports suggesting shared individual and structural determinants of infection between influenza and SARS-CoV-2. In the United States, there is evidence of higher COVID-19 incidence, morbidity and mortality among African American communities secondary to underlying inequities including structural racism [21]. These racial disparities were also observed during the 2009 H1N1 influenza pandemic with similar drivers including higher risk of exposure due to inability to social distance, higher burden of comorbidities and limited access to health care among African American and other racialized communities [22-24]. Similar early trajectories of COVID-19 and 2009 H1N1 have been noted in South Africa with higher burdens initially among higher income communities reporting more travel with rapid epidemiologic transition to lower income communities [25]. Finally, increases in COVID-19 in late May and June in some areas of the Southern hemisphere including countries across South America, Sub-Saharan Africa, East Asia and the Pacific were consistent with the timing of the traditional influenza season [26]. Specifically, in South Africa, epidemic growth is consistent with the influenza season and with epicenters of COVID-19 appearing to be similar to areas with increased influenza transmission [27, 28].

When this novel coronavirus emerged in early 2020, regional and national forecasting exercises drew on emerging data on the trajectory of detected cases of COVID-19 in the epicenters of China and Italy to translate their reproductive rates into best- and worst-case scenarios for populations all over the world. However, despite early comparison of the average mortality of influenza and SARS-CoV-2, there has not been a systematic evaluation of whether historical distributions in influenza burden are relevant in understanding the burden and distribution of SARS-CoV-2 infection. Descriptive epidemiological assessment within and across both influenza and COVID-19 may provide early insight into overlapping individual, structural, and environmental determinants of infection which have implications for public health responses. Moreover, documenting consistencies in the burden of disease across both pathogens may facilitate the generation of testable hypotheses, facilitating future analytic comparisons as well as future pandemic preparedness. In this study, we investigated global influenza transmission patterns from 2017 onwards to describe shared trends with the epidemiology of COVID-19.

## Methods

### Data abstraction and assessment of eligibility

#### Influenza

We used publicly available data to characterize temporal patterns of influenza across seven regions between 2017 and 2019. Organization of countries into regions was based on World Bank classifications, namely: East Asia and the Pacific, Europe and Central Asia, Latin America and the Caribbean, Middle East and North Africa, North America, South Asia, and Sub-Saharan Africa. Within each region, we assessed country-specific weekly reported cases of influenza, including both Influenza A and B, from 1 January 2017 to 31 December 2019. Data were abstracted from FluNet, a global web-based tool developed by the World Health Organization for influenza virologic surveillance, which captures weekly laboratory confirmed influenza cases. FluNet aggregates data from the National Influenza Centres of the Global Influenza Surveillance and Response System and other national influenza reference laboratories. Since more temperate zones in the Northern and Southern Hemispheres have generally opposing seasonal influenza trends, we report influenza case counts by calendar year from January 1 to December 31, which also facilitates comparisons with COVID-19. Finally, influenza case counts for the calendar year of 2020 were excluded given that public health measures focused on COVID-19 prevention and mitigation likely significantly decreased seasonal influenza cases for this time period [29]. The suspension of FluView, the core influenza reporting system in the United States, further precluded the inclusion 2020 influenza case counts.

We assessed patterns of missing data for each country and by year to determine eligibility for inclusion in analyses. For each year, countries where ≥ 90% of available weekly case reports were complete (assessed as the total number of non-missing records over the total number of all included weeks for that year) were included in analyses. For countries where data were <90% complete, we made the following determination: Countries where data missingness appeared to follow a seasonal pattern (e.g, data were missing only for weeks outside of the known influenza season, likely reflecting country-specific seasonal influenza reporting guidelines) were retained in analyses. Countries where weekly case counts were <90% complete and did not appear to follow a seasonal pattern, were denoted as missing.

### COVID-19

We abstracted the daily number of COVID-19 cases reported between 1 January 2020 and 31 October 2020 for all countries and corresponding World Bank regions using data from the World Health Organization Coronavirus Situation Report. As we are in the first year of COVID-19, it was not possible to assess missingness of data similar to the methods used for influenza.

### Regional Influenza and COVID-19 burden

#### Influenza

We assessed the overall burden of influenza for all included countries across and within each region. To do so, we summed the absolute number of weekly influenza cases for each region, country and year (January 1 to December 31 of each year) given the temporal variability in influenza seasons around the world. Countries were ranked in descending order within each region from highest to lowest number of absolute cases overall and for 2017, 2018, and 2019.

#### COVID-19

We assessed, and similarly ranked, in descending order, cumulative cases of COVID-19 as reported through 31 October within each region.

#### Comparison of regional burden of Influenza and COVID-19

We used regional plots for both influenza and COVID-19 cases to visualize cross-pathogen comparisons. For each region, we plotted the mean number of weekly influenza cases for the five highest ranked countries across 2017, 2018, and 2019. Influenza cases were averaged to ensure observed trends were robust to one-off spikes or changes to reporting systems in select years. Daily COVID-19 cases for the five highest ranked countries within each region were plotted using a 7-day moving average. We similarly plotted influenza and COVID-19 cases using per-capita proportions for these countries to facilitate standardized comparisons across countries and regions. Plots were visually inspected to confirm temporal patterns, and we described similarities or differences across and within all regions.

#### Within-country comparison of Influenza and COVID-19

We used plots to visually compare within-country trends for both influenza and COVID-19 over time. We assessed the top 20 countries with the highest burden of COVID-19 for each region, further restricting plots to those countries where influenza data were eligible for inclusion. As with regional comparisons, we plotted the mean number of weekly influenza cases across 2017, 2018, and 2019 and daily COVID-19 cases were plotted using a 7-day moving average.

## Results

Influenza cases were assessed for 171 countries. For each year, more than half (ranging from 55.6%-56.7%) of all countries reported ≥90% complete data (**Multimedia Appendix 1**). The proportion of countries where data were <90% complete but followed a pattern of seasonal missingness were also included in influenza analyses, ranged from 22.8% in 2017 to 24.0% in 2019. Differences in data completion and patterns of missingness were evident across all regions. The overall influenza burden between 2017 and 2019 varied by region and by countries within all seven identified regions (**Table 1**). Regions with the highest absolute burden of influenza included North America (903,554 cases), Europe and Central Asia (599,820 cases), and East Asia and the Pacific (389,657 cases). Countries with the highest proportion of regional influenza cases were as follows: East Asia and the Pacific (China; 77.5%); Europe and Central Asia (the United Kingdom; 17.7%), Latin America and the Caribbean (Mexico, 20.2%), Middle East and North Africa (Qatar, 40.5%), North America (the United States, 82.9%), South Asia (India, 46.6%), and Sub-Saharan Africa (South Africa, 18.7%). Across regions, countries with the highest overall burden of influenza from 2017 to 2019, included the United States (749,472 cases), China (302,126 cases), Canada (154,082 cases), the United Kingdom (106,087 cases), and Norway (71,727 cases).

**TABLE 1.**
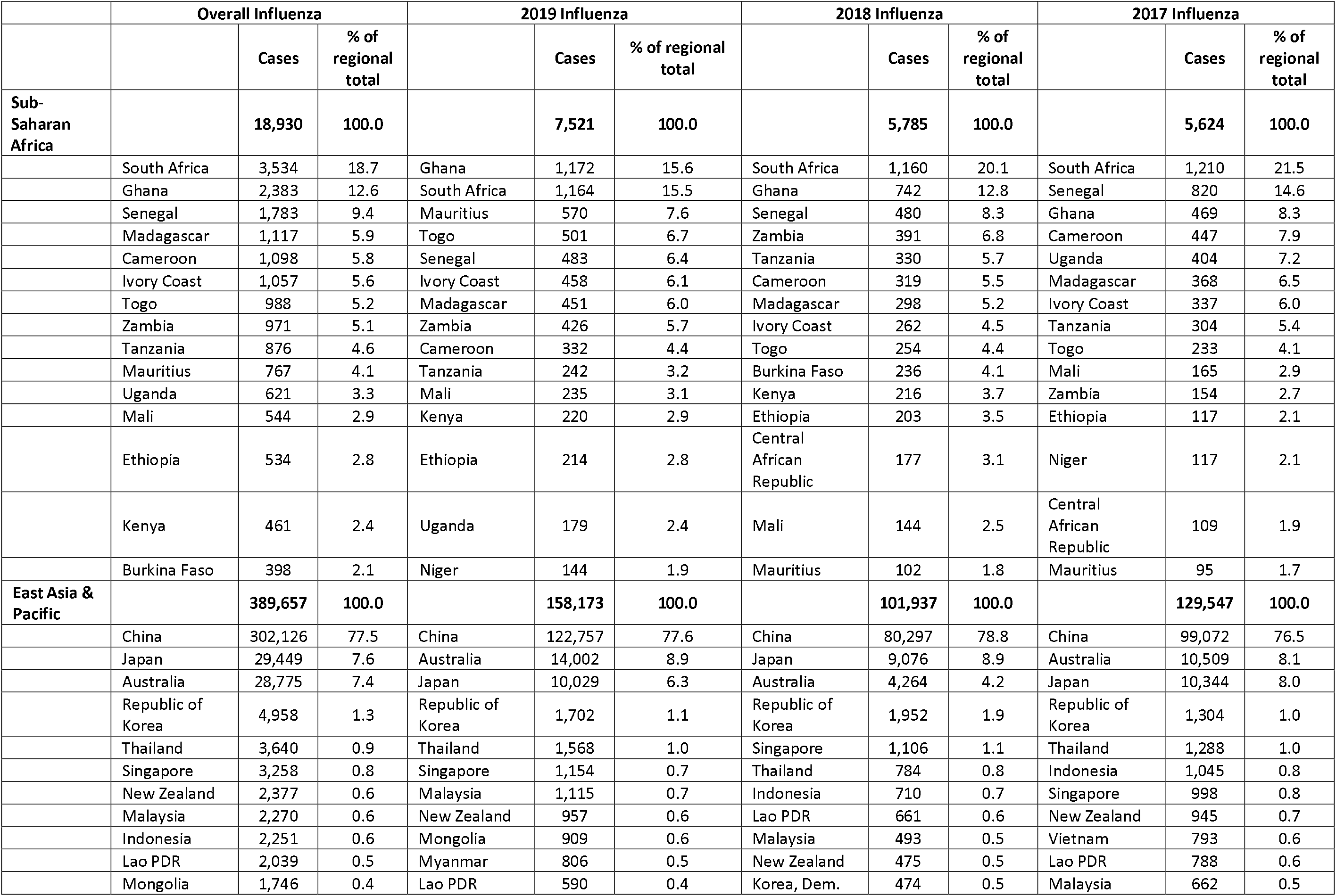
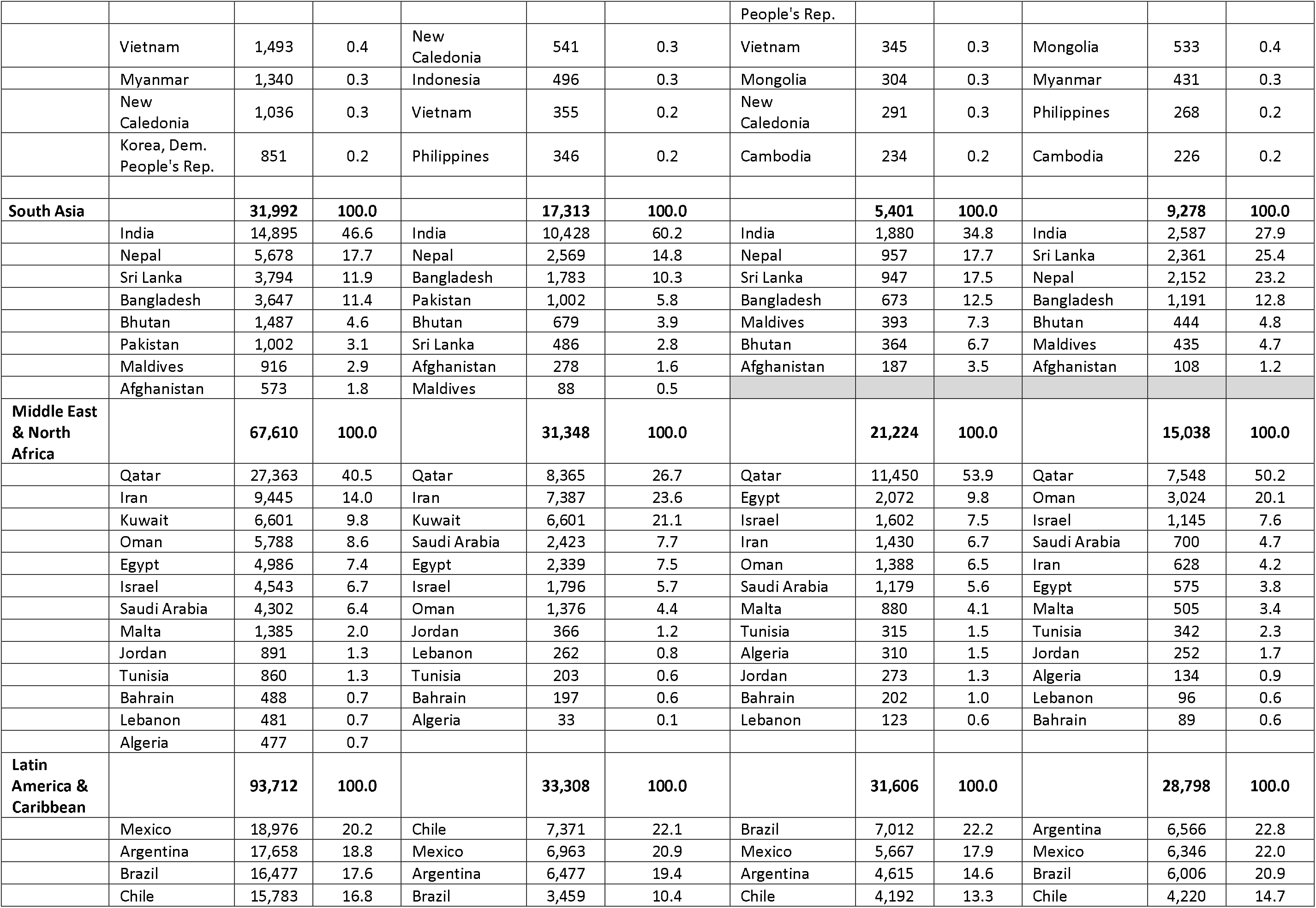
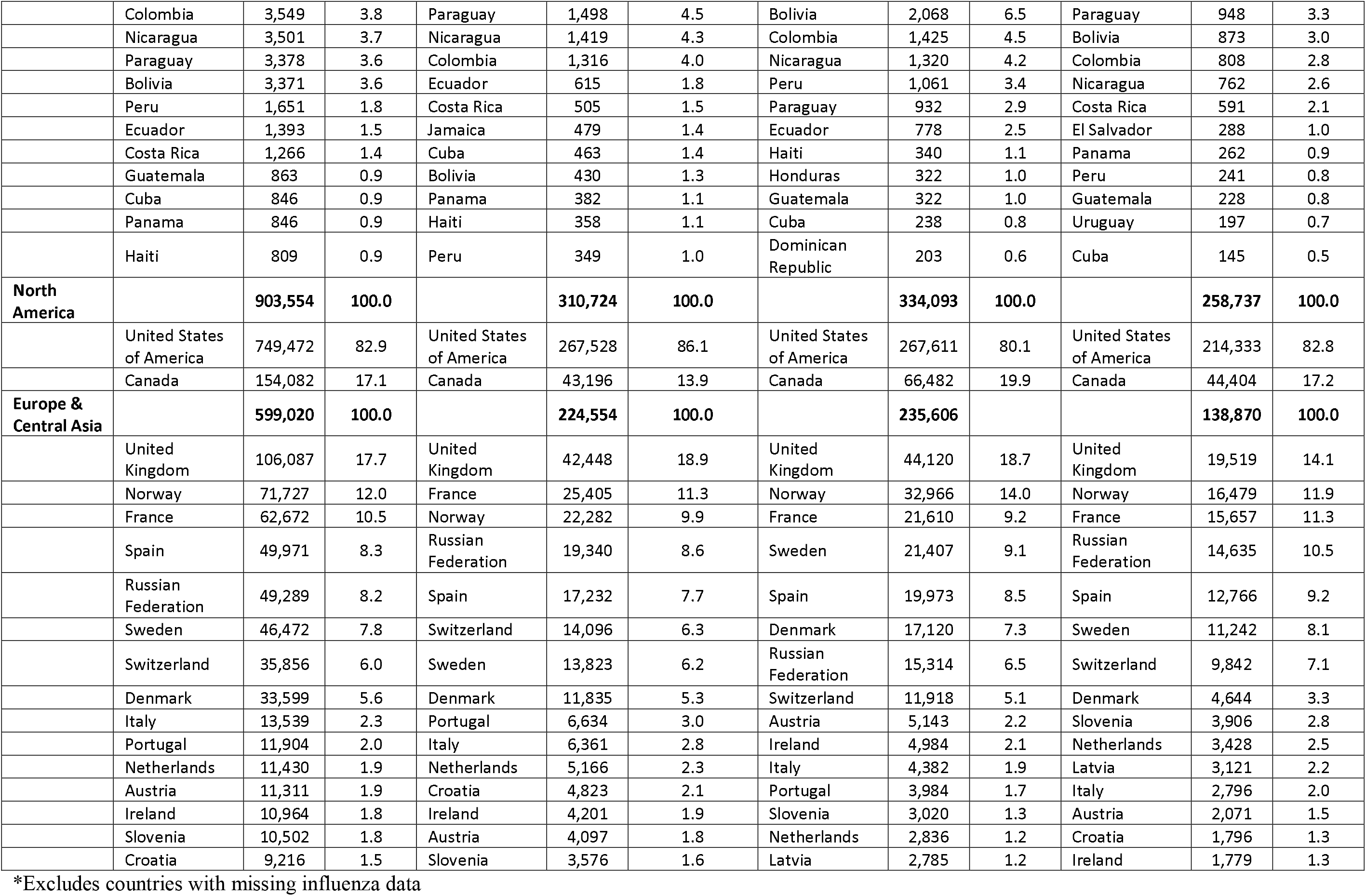
Influenza cases for top 15 countries by World Bank region, overall and by year from January 1 2017 to December 31 2019^*^

Regions with the highest absolute burden of COVID-19 included Latin America and the Caribbean (11,045,364 cases), Europe and Central Asia (9,197,241 cases), South Asia (9,107,653 cases), and North America (9,081,471 cases) (**Table 2**). Within each region, countries with the highest proportion of COVID-19 regional cases were as follows: East Asia and the Pacific (Indonesia; 34.2%); Europe and Central Asia (Russia; 17.6%), Latin America and the Caribbean (Brazil, 49.7%), Middle East and North Africa (Iran, 22.7%), North America (the United States, 97.5%), South Asia (India, 89.3%), and Sub-Saharan Africa (South Africa, 63.5%) (**Table 2)**. Specific countries with the highest overall burden of COVID-19 cases as of October 31, 2020 included the United States (8,852,730 cases), India, (8,137,119 cases), Brazil (5,494,376 cases), Russia (1,618,116 cases), and France (1,299,278 cases) (**Table 2)**.

**TABLE 2.** COVID-19 cases for top 15 countries by World Bank region, from January 1 2020 to October 31 2020

|  |  | COVID-19 cases | % of regional total |
| --- | --- | --- | --- |
| **Sub-Saharan Africa** |  | **1,139,329** | **100.0** |
|  | South Africa | 723,682 | 63.5 |
|  | Ethiopia | 95,789 | 8.4 |
|  | Nigeria | 62,691 | 5.5 |
|  | Kenya | 53,797 | 4.7 |
|  | Ghana | 48,055 | 4.2 |
|  | Cameroon | 21,793 | 1.9 |
|  | Cote d'Ivoire | 20,628 | 1.8 |
|  | Madagascar | 16,968 | 1.5 |
|  | Zambia | 16,415 | 1.4 |
|  | Senegal | 15,593 | 1.4 |
|  | Sudan | 13,804 | 1.2 |
|  | Namibia | 12,907 | 1.1 |
|  | Mozambique | 12,777 | 1.1 |
|  | Uganda | 12,410 | 1.1 |
|  | Guinea | 12,020 | 1.1 |
| **East Asia & Pacific** |  | **1,191,632** | **100.0** |
|  | Indonesia | 406,945 | 34.2 |
|  | Philippines | 378,933 | 31.8 |
|  | Japan | 100,392 | 8.4 |
|  | China | 91,893 | 7.7 |
|  | Singapore | 58,003 | 4.9 |
|  | Myanmar | 51,496 | 4.3 |
|  | Malaysia | 30,889 | 2.6 |
|  | Australia | 27,582 | 2.3 |
|  | Korea, Rep. | 26,511 | 2.2 |
|  | French Polynesia | 7,262 | 0.6 |
|  | Guam | 4,579 | 0.4 |
|  | Thailand | 3,780 | 0.3 |
|  | New Zealand | 1,601 | 0.1 |
|  | Vietnam | 1,177 | 0.1 |
|  | Papua New Guinea | 589 | 0.0 |
| **South Asia** |  | **9,107,653** | **100.0** |
|  | India | 8,137,119 | 89.3 |
|  | Bangladesh | 406,364 | 4.5 |
|  | Pakistan | 332,186 | 3.6 |
|  | Nepal | 168,235 | 1.8 |
|  | Afghanistan | 41,334 | 0.5 |
|  | Maldives | 11,643 | 0.1 |
|  | Sri Lanka | 10,424 | 0.1 |
|  | Bhutan | 348 | 0.0 |
| **Middle East & North Africa** |  | **2,662,465** | **100.0** |
|  | Iran | 604,952 | 22.7 |
|  | Iraq | 470,633 | 17.7 |
|  | Saudi Arabia | 346,880 | 13.0 |
|  | Morocco | 215,294 | 8.1 |
|  | Qatar | 132,343 | 5.0 |
|  | United Arab Emirates | 131,508 | 4.9 |
|  | Kuwait | 125,337 | 4.7 |
|  | Oman | 114,434 | 4.3 |
|  | Egypt | 107,385 | 4.0 |
|  | Bahrain | 81,466 | 3.1 |
|  | Lebanon | 79,529 | 3.0 |
|  | Jordan | 69,306 | 2.6 |
|  | West Bank and Gaza | 64,741 | 2.4 |
|  | Libya | 60,628 | 2.3 |
|  | Tunisia | 58,029 | 2.2 |
| **Latin America & Caribbean** |  | **11,045,364** | **100.0** |
|  | Brazil | 5,494,376 | 49.7 |
|  | Argentina | 1,143,800 | 10.4 |
|  | Colombia | 1,053,122 | 9.5 |
|  | Mexico | 912,811 | 8.3 |
|  | Peru | 897,594 | 8.1 |
|  | Chile | 508,571 | 4.6 |
|  | Ecuador | 167,147 | 1.5 |
|  | Bolivia | 141,484 | 1.3 |
|  | Panama | 132,045 | 1.2 |
|  | Dominican Republic | 126,332 | 1.1 |
|  | Costa Rica | 107,570 | 1.0 |
|  | Guatemala | 107,339 | 1.0 |
|  | Honduras | 96,150 | 0.9 |
|  | Venezuela | 91,280 | 0.8 |
|  | Puerto Rico | 65,743 | 0.2 |
| **North America** |  | **2,662,465** | **100.0** |
|  | United States of America | 8,852,730 | 97.5 |
|  | Canada | 228,542 | 2.5 |
|  | Bermuda | 199 | 0.0 |
| **Europe & Central Asia** |  | **9,197,241** | **100.0** |
|  | Russian Federation | 1,618,116 | 17.6 |
|  | France | 1,299,278 | 14.1 |
|  | Spain | 1,212,190 | 13.2 |
|  | United Kingdom | 989,749 | 10.8 |
|  | Italy | 647,674 | 7.0 |
|  | Germany | 518,753 | 5.6 |
|  | Belgium | 441,807 | 4.8 |
|  | Ukraine | 387,481 | 4.2 |
|  | Turkey | 373,154 | 4.1 |
|  | Netherlands | 340,974 | 3.7 |
|  | Poland | 340,834 | 3.7 |
|  | Czech Republic | 323,673 | 3.5 |
|  | Israel | 314,244 | 3.4 |
|  | Romania | 235,586 | 2.6 |
|  | Switzerland | 153,728 | 1.7 |

Seasonal variation in influenza cases was observed within all regions. Seasonal trends were most pronounced across countries for North America, Europe and Central Asia, Middle East and North Africa, East Asia and the Pacific, and Latin America and the Caribbean (**Figure 1)**. Seasonal trends were also evident but largely inconsistent across countries within South Asia and sub-Saharan Africa. These dynamics were mainly present when trends were assessed by per capita proportions instead of case counts (**Figure 2**).

**Figure 1.**
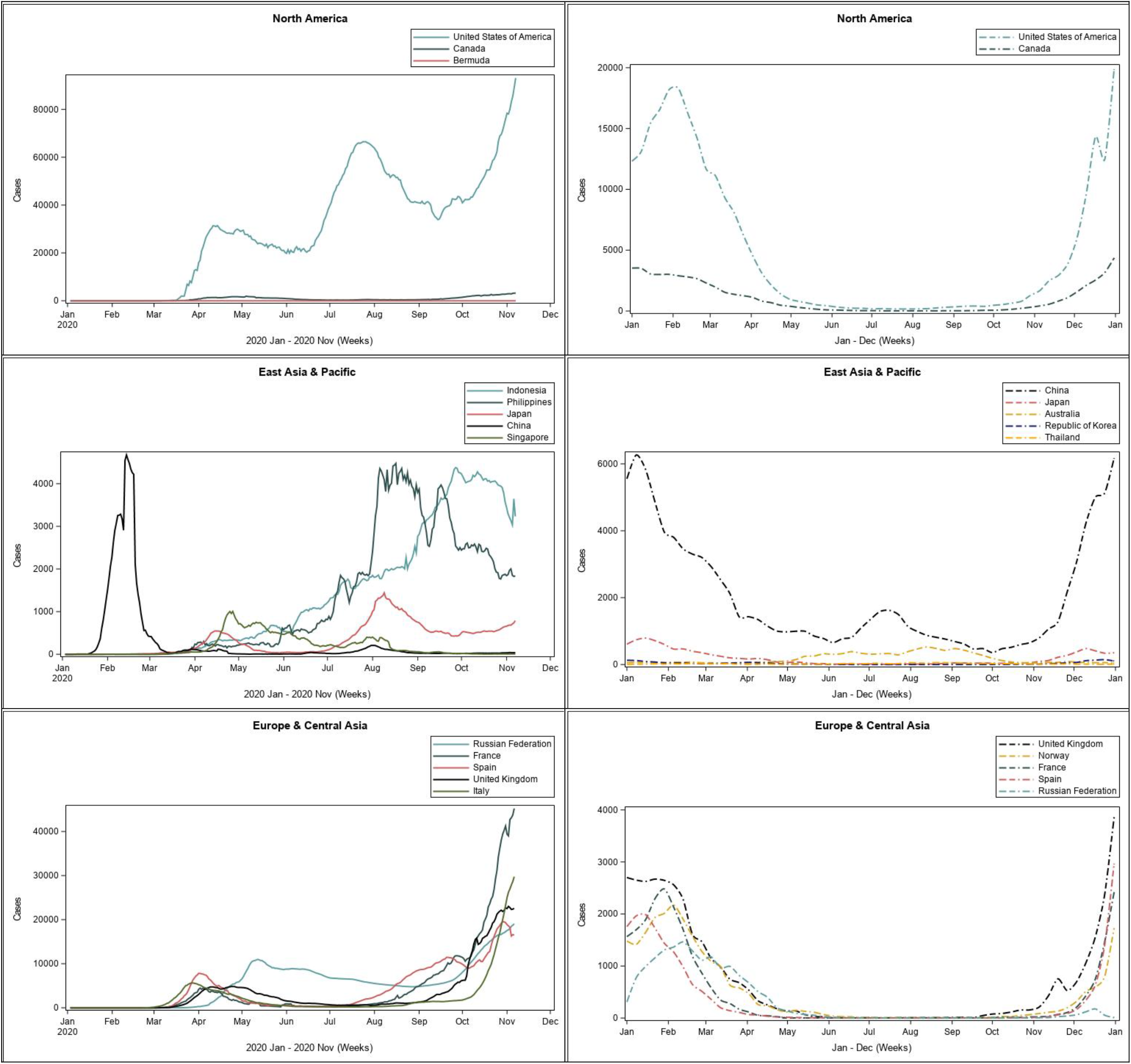
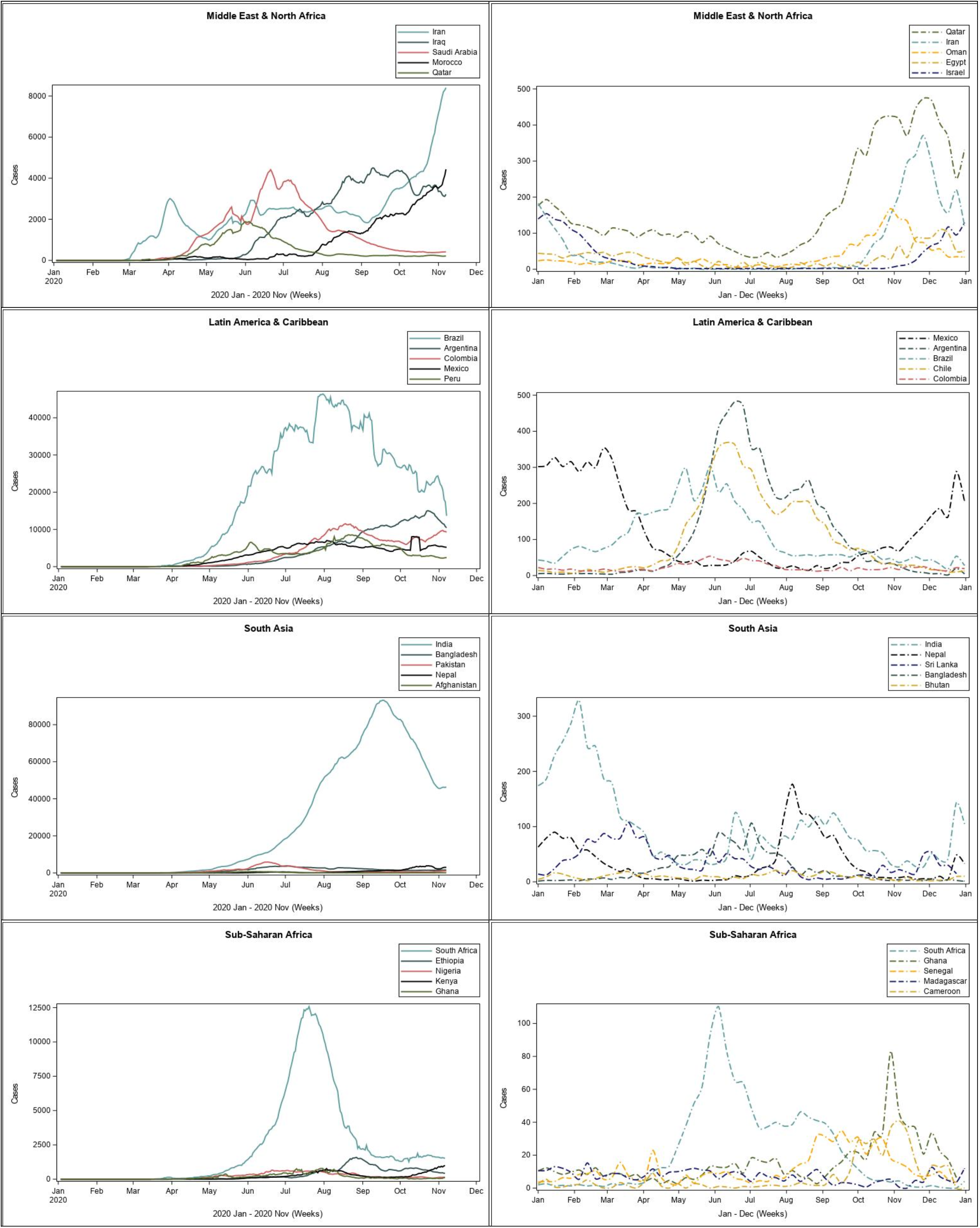
Weekly trends in new COVID-19 cases (left panel, solid lines) and Influenza cases (right panel, dashed lines) presented by World Bank region. Daily COVID-19 cases were extracted from WHO and are presented as a 7-day moving average between January 1 2020 and October 31 2020. Weekly influenza cases were extracted from FluNet and averaged by week across all reporting years (2017-2019).

**Figure 2.**
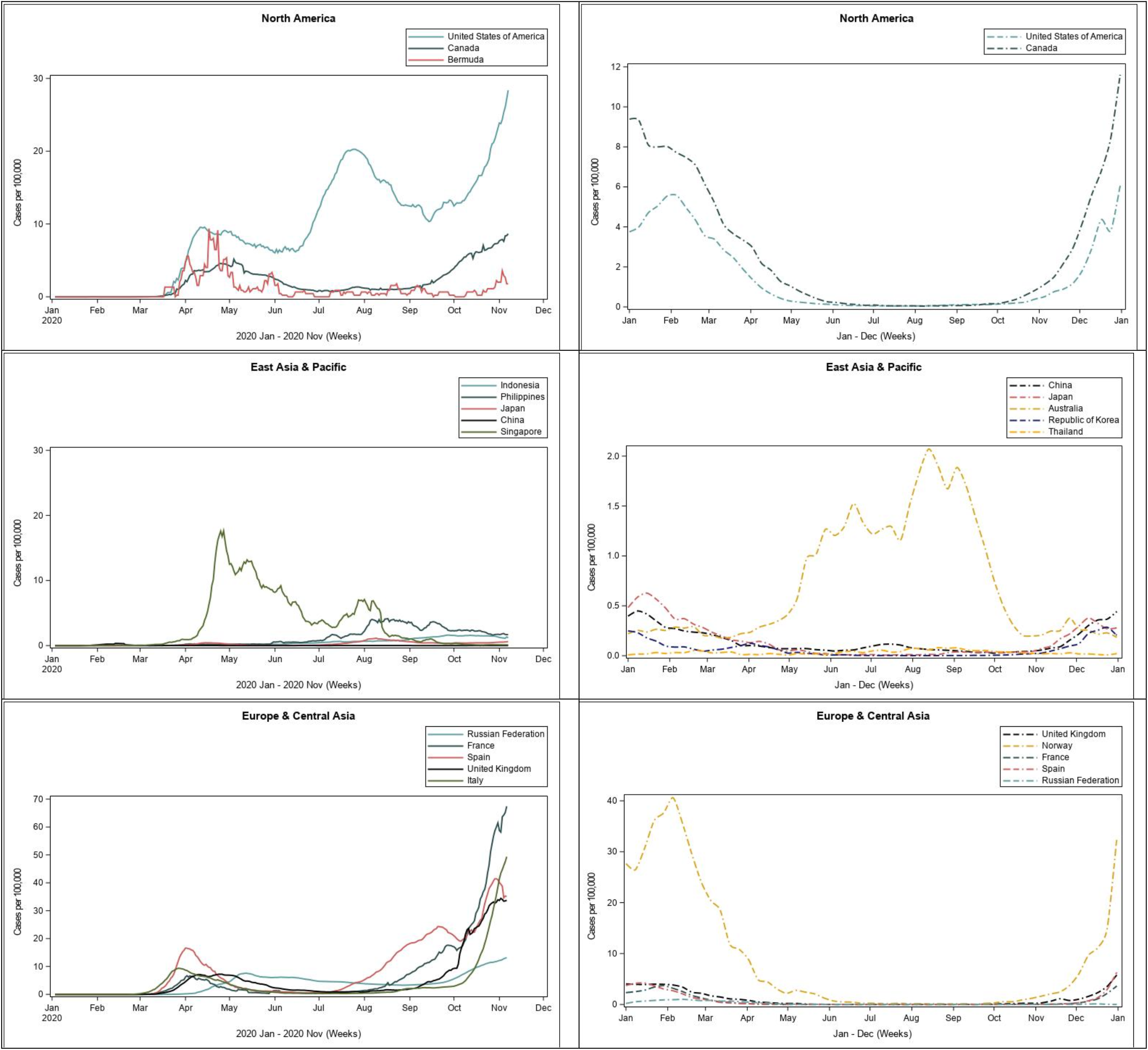
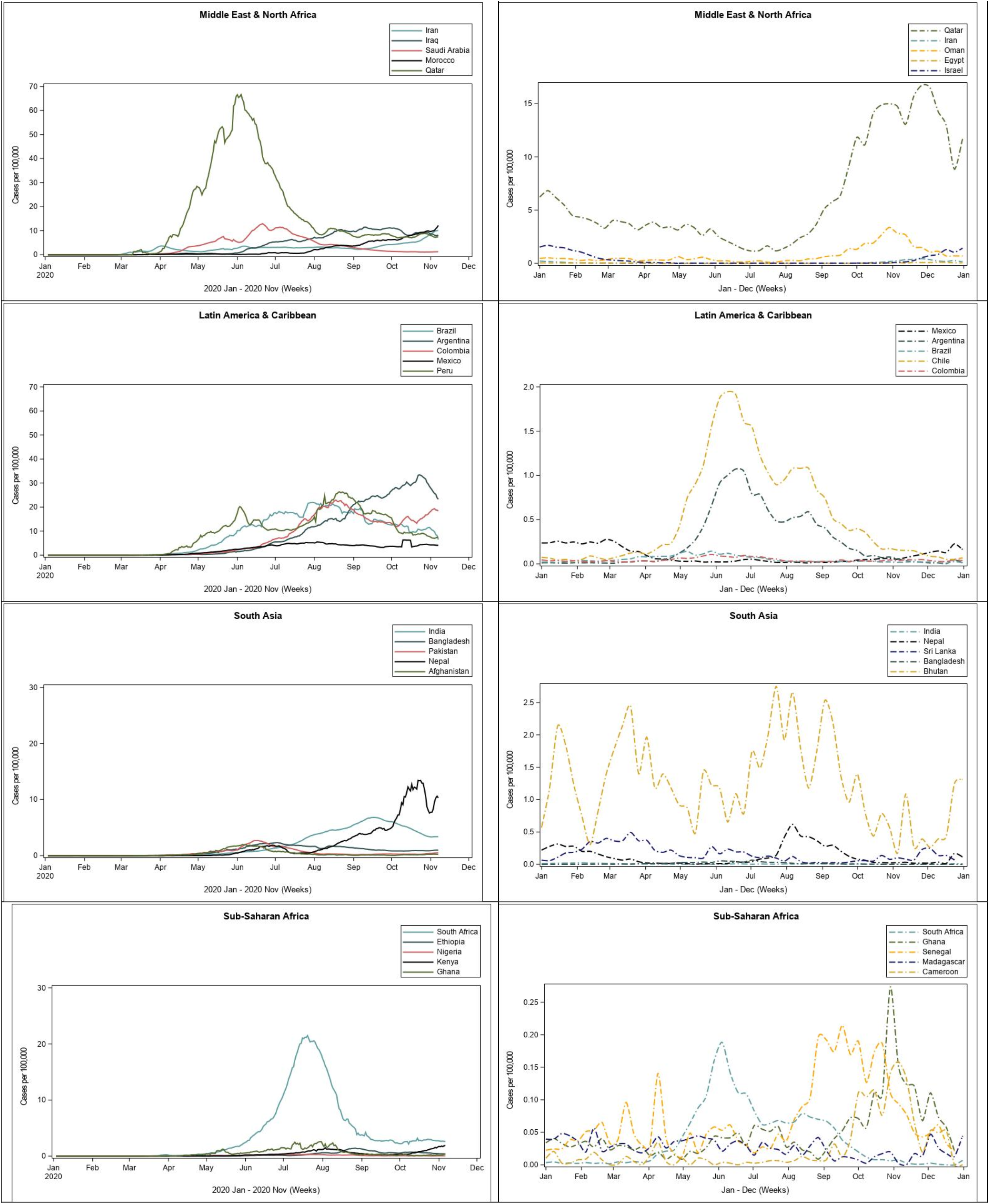
Weekly trends in new COVID-19 cases per capita (left panel, solid lines) and Influenza cases per capita (right panel, dashed lines) presented by World Bank region. Daily COVID-19 cases were extracted from WHO and are presented as a 7-day moving average, per 100,000 persons, between January 1 2020 and October 31 2020. Weekly influenza cases were extracted from FluNet and averaged by week, per 100,000 persons, across all reporting years (2017-2019).

COVID-19 epidemics generally followed trends similar to those observed for influenza from 2017-2019 in the regions with the most pronounced influenza seasonal trends (**Figure 2**). This was more apparent in Europe and Central Asia with an early wave of COVID-19 epidemics in several countries from March – May followed by significant drops in transmission in the summer and a second wave in the fall. With the exception of the United States that has seen sustained transmission over the year, Canada and Bermuda, have also had COVID-19 epidemics largely similar to those observed in European countries. In Latin America and the Caribbean, COVID-19 epidemics in Brazil, Argentina, Peru, Colombia peaked during the summer and started decreasing in fall corresponding to influenza trends in the region. Countries in sub-Saharan Africa, South Asia, and Middle East and North Africa have shown more heterogeneity in their COVID-19 epidemics.

Similarities between COVID-19 and influenza trends were more marked within individual countries irrespective of region (**Multimedia Appendix 2**). Peaks of COVID-19 epidemics in exemplary countries, including Australia, Brazil, India, South Asia and South Africa, coincided with months of peak influenza activity in those countries from 2017-2019 (**Figure 3**).

**Figure 3.**
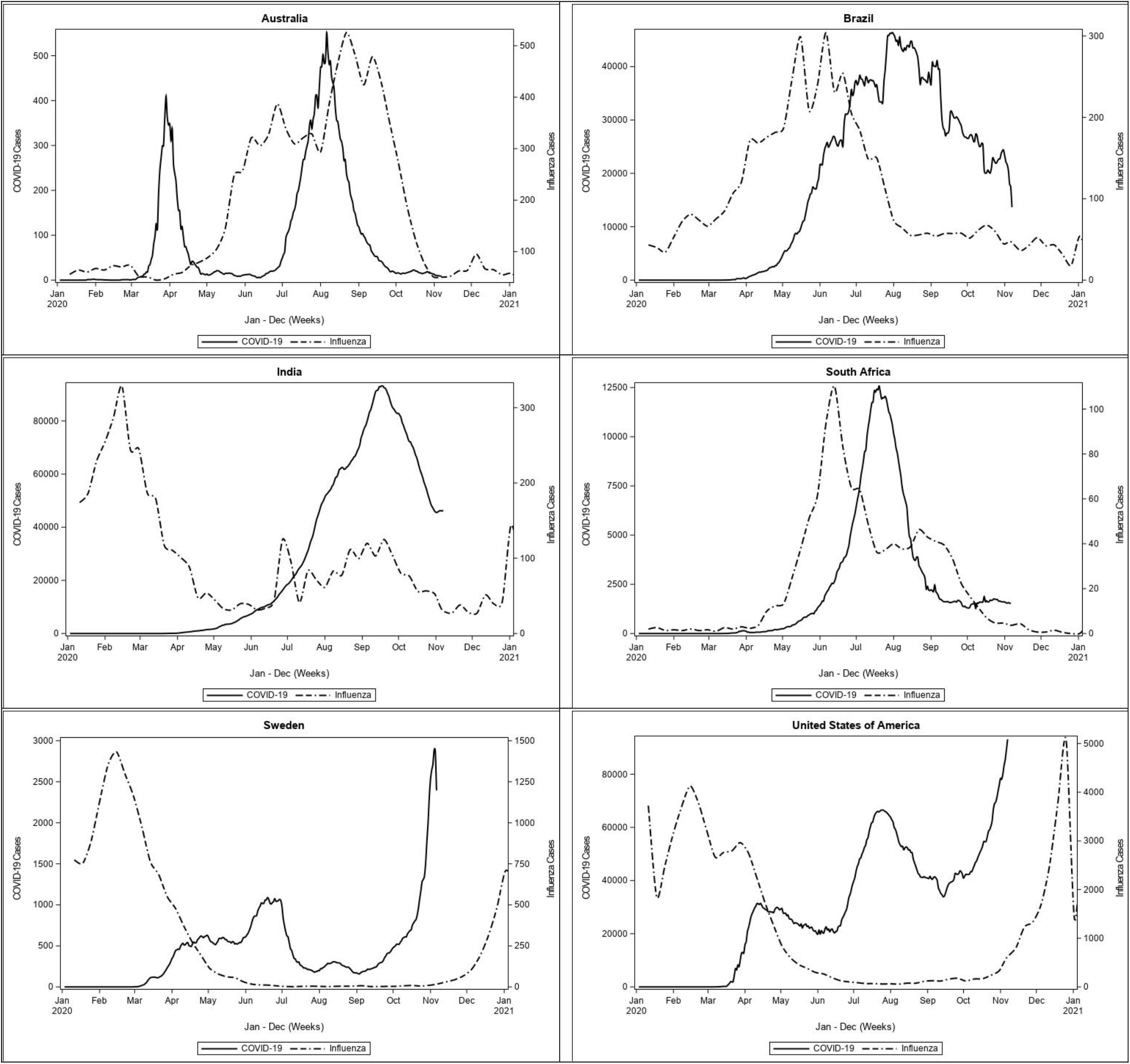
Weekly trends in new COVID-19 cases (solid lines) and Influenza cases (dashed lines) presented by country. Daily COVID-19 cases were extracted from WHO and are presented as a 7-day moving average between January 1 2020 and October 31 2020. Weekly influenza cases were extracted from FluNet and averaged by week across all reporting years (2017-2019).

## Discussion

In this descriptive epidemiological study, we found ecological consistency between the seasonal influenza trends from 2017-2019 and the COVID-19 trends seen to date. Specifically, this similarity was highest in regions with more pronounced influenza seasonality including North America, Europe and Central Asia and Latin America and the Caribbean regions where peaks of COVID-19 cases coincided with periods of high influenza activity. In East Asia and the Pacific and Sub-Saharan Africa, there was more marked heterogeneity in COVID-19 pandemics in countries of these regions. However, within-country comparisons of 2017-2019 influenza epidemics and COVID-19 cases were evident irrespective of region. The ecological consistency in the seasonal trends of COVID-19 and the recent burden of influenza is suggestive of likely shared individual, structural, and environmental determinants of infection which has implications for public health.

Outside of public health interventions, there is generally less consensus on the determinants that potentiate or inhibit COVID-19 transmission. However, given decades of study, there is more information available on the individual, population-level, and environmental determinants of influenza. At the individual level, underlying immunity is related to previous exposure to similar strains and vaccine uptake and effectiveness. Effectiveness of vaccines depends on circulating influenza strains i.e. pandemic and non-pandemic, or seasonal, secondary to antigenic drift and shift of influenza viruses. Population-level characteristics include vaccine coverage and shield immunity, indoor air quality, population density, social norms, and health care infrastructure [30-32]. Finally, environmental factors affecting influenza include temperature, air pollution, and humidity [30, 33]. Given how rapidly COVID-19 has emerged, conclusions of different studies exploring the role of environmental conditions have differed significantly. One study found no associations between latitude and temperature with COVID-19 epidemic growth using a two-week exposure period in March across 144 geopolitical areas [34]. However, a “living” review leveraging data from December, 2019 onwards from nearly 4000 locations across the world showed significant differences in COVID-19 burden associated with temperature, humidity, pollution, and other environmental factors [35]. Collectively, these studies suggest the potential for at least some seasonal variation secondary to changes in environmental conditions in the transmission of SARS-CoV-2 similar to other respiratory pathogens, including influenza and other seasonal coronaviruses [36-39].

Influenza seasonality is more pronounced in more temperate areas of the Northern and Southern hemispheres consistent with opposing fall and winter seasons. A recent assessment of the seasonality of infectious pathogens included individual-level determinants such as growing immunity throughout an influenza season secondary to vaccination or exposure, that results in achieving effective levels of shield immunity limiting transmission [40]. There has been some study of existing immunity to COVID-19 mediated via cross-reactivity with other coronaviruses [40, 41]. However, lesser population-wide immunity could result in sustained transmission even during times with less transmission of other respiratory pathogens. Influenza seasonality may also be secondary to population-level changes, including potentially higher crowding in colder weather, increased use of respite care and housing, people returning to work and school, and increased use of transit services that may collectively facilitate transmission. Environmental conditions may further affect individual susceptibility to infection as well as contact or respiratory droplet transmission patterns. While there is likely a relationship between testing infrastructure and influenza reporting, the consistency in the COVID-19 and influenza trends among countries within each region was notable. COVID-19 emerged in East Asia and the Pacific though there was relatively limited COVID-19 spread in several settings in March and April, 2020 potentially secondary to widely implemented non-pharmacologic interventions [29]. Notably, and consistent with influenza season, there was a resurgence of COVID-19 in Australia starting in July and ending in September, 2020 [42-44].

A hallmark of COVID-19 has been the role of social determinants of health with socioeconomic status, living conditions, access to health care, and underlying comorbidities being consistent predictors of COVID-19 incidence and mortality around the world [45, 46]. Moreover, structural factors including racism have been documented in several settings including the United States, United Kingdom, Sweden, and Canada, but likely relevant in many more settings not reporting on racial identity of those affected [47-49]. Notably, similar disparities were observed in the 2009 H1N1 pandemic in the United States though there has been limited documentation of socioeconomic disparities of influenza in most settings [22, 50]. While disparities are well documented for other infectious diseases such as HIV, these disparities evolve across cycles of transmission spanning years. Given how rapidly disparities emerged with COVID-19, interventions to address disparities may be observable over shorter time horizons and provide insight into effective interventions to address pervasive social health inequities. Secondary to intersections of differential access to health care and medical mistrust, the development of a generally effective vaccine has not mitigated disparities [51-53]. Leveraging frameworks focused on optimizing implementation of influenza vaccination strategies including increased equity will be critical in informing SARS-CoV-2 vaccines when available [54, 55]. Moreover, there has been significant research into studying correlates of protection for influenza vaccine development as well as the role of cellular immunity in complementing the humoral response which are both actively being studied for COVID-19 [56-59]. Finally, COVID-19 is rapidly increasing our understanding of post-viral morbidities including chronic fatigue syndrome and chronic myofascial pain syndromes as well as potential cardiovascular complications [60]. While post-viral complications have been described for other viruses, the known exposure to COVID-19 as an etiological agent may provide further insight into the mechanisms and potential interventions [61-66].

This study has several limitations. Risk of ecological fallacy is relevant where individuals in generally lower burden regions or countries may have high risks of acquisition and transmission secondary to micro-epidemics, including in those in congregate living settings, refugee and migrant work camps, long term care facilities, homeless shelters, and prisons. Missingness in data aggregated by FluNet is another limitation. We assumed that countries where influenza data was missing in a seasonal pattern was a reflection of very few case counts and/or reduced reporting outside of the typical influenza season, and indeed seasonal “missingness” was most apparent for countries in the European region, where the influenza season has been well documented. However, by excluding countries with > 10% data missingness that did not follow a seasonal influenza pattern, several countries in the Pacific and in sub-Saharan Africa (e.g. Sudan) and were not included in the comparison of rankings despite high COVID-19 and influenza burden. There have been examples of COVID-19 resurgence outside of traditional influenza season such as in South Africa potentially due to the emergence of new variants of COVID-19 highlighting the complex intersection of drivers of transmission in specific subnational areas [67]. Finally, there have been dramatic decreases in the burden of influenza in the 2020 season likely due to an intersection of the effects of non-pharmacological interventions combined with decreases in international travel. It is unclear how this will affect vulnerability to future seasons of influenza [68].

In addition, absolute influenza and COVID-19 case counts were reported, rather than positivity rates, assuming that biases present in interpreting positivity rates (e.g. laboratory infrastructure to implement broad testing strategies) is likely reflected in the total number of cases, across both influenza and COVID-19. Heterogeneity in testing capacity, test kit availability, and in guidelines for prioritizing testing eligibility for COVID-19, may additionally challenge direct comparisons between COVID-19 and influenza case counts across countries. Testing capacity for COVID-19 has been highly dynamic, with continued limited capacity to meet screening and testing demands still evident in some countries [69]. In the case of influenza, country-wide variations in diagnostic testing strategies may belie differences in case detection. For instance, rapid influenza diagnostic tests are commonly used in clinical settings in many countries yet have documented lower sensitivity relative to other diagnostic methods [70].

Relatedly, contact tracing strategies for COVID-19 may vary across and within countries. As the pandemic has progressed, strategies have adapted to testing individuals with exposure to confirmed cases, irrespective of their symptomology [71]. This approach is not typically used for influenza. Finally, evidence suggests that non-pharmaceutical interventions for COVID-19 may concurrently limit influenza spread, due to their shared modes of transmission [72, 73]. While COVID-19 transmission has likely persisted due to unknown baseline population immunity, limited influenza detection may reflect decreased propensity for testing outside of the influenza season in some countries, as previously highlighted. Nonetheless, these data further underscore the potential utility to leverage our understanding and mitigation strategies for other respiratory pathogens to understand and combat COVID-19 and future pandemics due to respiratory pathogens.

The results presented here suggest shared regional differential distribution in the global burden of COVID-19 with that of influenza. This ecological overlap suggests that the underlying dynamics of transmission of influenza and COVID-19 are similar as they relate to spatial, seasonal, population structure and the disparities that lead to disproportionate vulnerabilities in various population groups. In considering the host-pathogen-environment framework commonly used for infectious diseases, emerging infections represent novel pathogens, but the populations and environment under which they emerge, and spread are the same. Similarly, the similarities observed here suggest the potential utility of informing COVID-19 prevention and mitigation interventions with the vast data available characterizing critical issues including the relative risks of indoor vs. outdoor transmission, risks of aerosolization for influenza, and even optimal delivery of vaccines and chemoprophylaxis and treatment strategies. Ultimately, regional and national public health systems preparing and responding to the current and future waves of COVID-19 may benefit from evaluating the relative burden of past respiratory pathogens and the impact of non-pharmacologic, pharmacologic, and prophylactic intervention strategies as prior information when forecasting and implementing interventions, and when evaluating the transmission impact of interventions applied to date in the context of potential seasonal effects on transmission.

## Supporting information

Multimedia Appendix 1

Multimedia Appendix 2

## Data Availability

All data reported in this manuscript are publicly available. Influenza data were abstracted from FluNet, a global web-based tool developed by the World Health Organization for influenza virological surveillance, which captures weekly laboratory confirmed influenza cases. COVID-19 data through 31 May, 2020 were abstracted from the World Health Organization Coronavirus Situation Report.

https://www.who.int/influenza/gisrs_laboratory/flunet/en/

https://reliefweb.int/report/world/coronavirus-disease-covid-19-situation-report-132-31-may-2020

