## Supplementary material for "Descriptive Epidemiological Assessment of the Relationship between the Global Burden of Influenza from 2017-2019 and COVID-19": Multimedia Appendix 1

**SUPPLEMENTAL TABLE 1.** Categories of missingness for 171 countries reporting influenza cases between January 2017 and December 2019 extracted from FluNet, overall and by region

|  | **All regions** | | **Sub-Saharan Africa** | | **South Asia** | | **North America** | | **Middle East & North Africa** | | **Latin American & Caribbean** | | **Europe & Central Asia** | | **East Asia & Pacific** | |
| --- | --- | --- | --- | --- | --- | --- | --- | --- | --- | --- | --- | --- | --- | --- | --- | --- |
|  | N=171 | | N=34 | | N=8 | | N=4 | | N=18 | | N=34 | | N=49 | | N=20 | |
|  | **n** | **%** | **n** | **%** | **n** | **%** | **n** | **%** | **n** | **%** | **n** | **%** | **n** | **%** | **n** | **%** |
| **2017** |  |  |  |  |  |  |  |  |  |  |  |  |  |  |  |  |
| ≥ 90 Complete | 95 | 55.6 | 20 | 58.8 | 5 | 62.5 | 2 | 66.7 | 9 | 50.0 | 17 | 50.0 | 25 | 51.0 | 16 | 80.0 |
| Missing, seasonal | 39 | 22.8 | 6 | 17.6 | 2 | 25.0 | 0 | . | 3 | 16.7 | 2 | 5.9 | 21 | 42.9 | 3 | 15.0 |
| Missing, not seasonal | 37 | 21.6 | 8 | 23.5 | 1 | 12.5 | 1 | 33.3 | 6 | 33.3 | 15 | 44.1 | 3 | 6.1 | 1 | 5.0 |
|  | **n** | **%** | **n** | **%** | **n** | **%** | **n** | **%** | **n** | **%** | **n** | **%** | **n** | **%** | **n** | **%** |
| **2018** |  |  |  |  |  |  |  |  |  |  |  |  |  |  |  |  |
| ≥ 90 Complete | 97 | 56.7 | 20 | 58.8 | 5 | 62.5 | 2 | 66.7 | 8 | 44.4 | 19 | 55.9 | 26 | 53.1 | 16 | 80.0 |
| Missing, seasonal | 41 | 24.0 | 6 | 17.6 | 2 | 25.0 | 0 | . | 4 | 22.2 | 3 | 8.8 | 21 | 42.9 | 3 | 15.0 |
| Missing, not seasonal | 33 | 19.3 | 8 | 23.5 | 1 | 12.5 | 1 | 33.3 | 6 | 33.3 | 12 | 35.3 | 2 | 4.1 | 1 | 5.0 |
|  | **n** | **%** | **n** | **%** | **n** | **%** | **n** | **%** | **n** | **%** | **n** | **%** | **n** | **%** | **n** | **%** |
| **2019** |  |  |  |  |  |  |  |  |  |  |  |  |  |  |  |  |
| ≥ 90 Complete | 96 | 56.1 | 20 | 58.8 | 6 | 75.0 | 2 | 66.7 | 9 | 50.0 | 18 | 52.9 | 24 | 49.0 | 16 | 80.0 |
| Missing, seasonal | 41 | 24.0 | 6 | 17.6 | 2 | 25.0 | 0 | . | 3 | 16.7 | 3 | 8.8 | 22 | 44.9 | 3 | 15.0 |
| Missing, not seasonal | 34 | 19.9 | 8 | 23.5 | 0 | . | 1 | 33.3 | 6 | 33.3 | 13 | 38.2 | 3 | 6.1 | 1 | 5.0 |
