## Supplementary figures and images for "Descriptive Epidemiological Assessment of the Relationship between the Global Burden of Influenza from 2017-2019 and COVID-19"

### Afghanistan1.jpeg

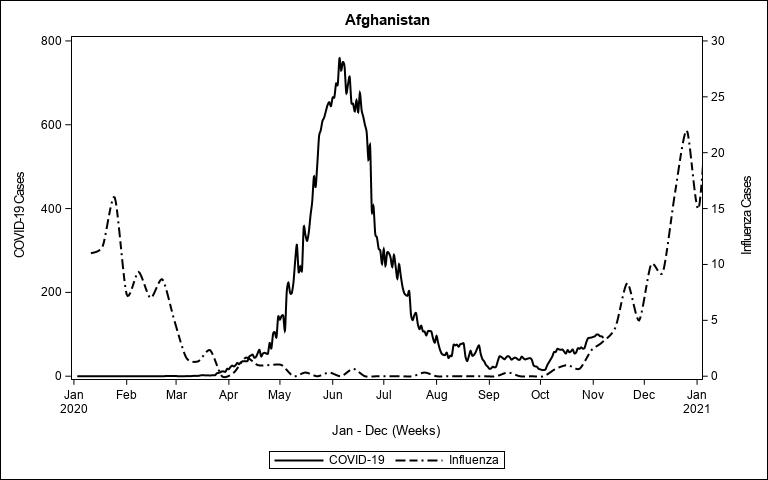

### Algeria1.jpeg

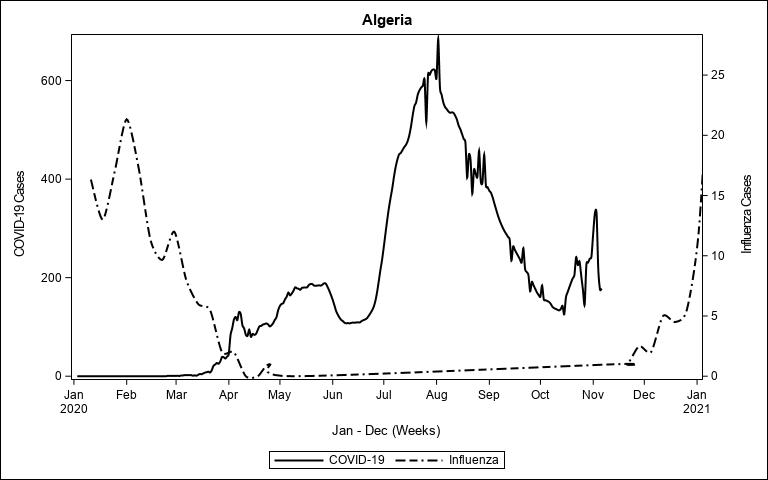

### Argentina1.jpeg

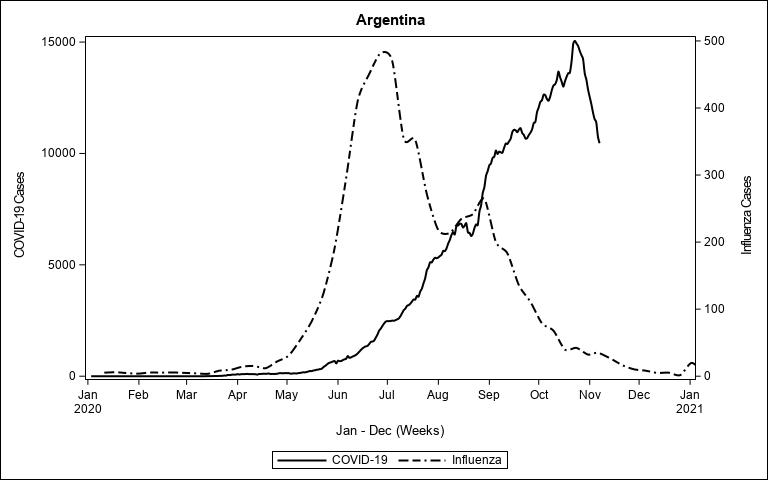

### Australia3.jpeg

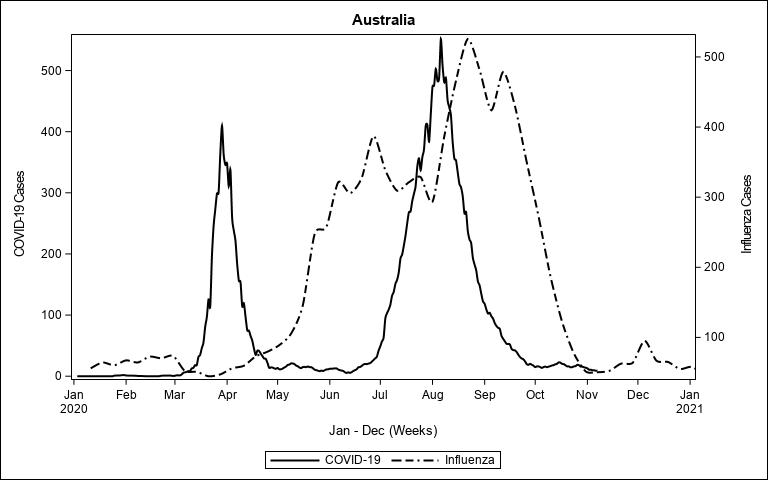

### Austria1.jpeg

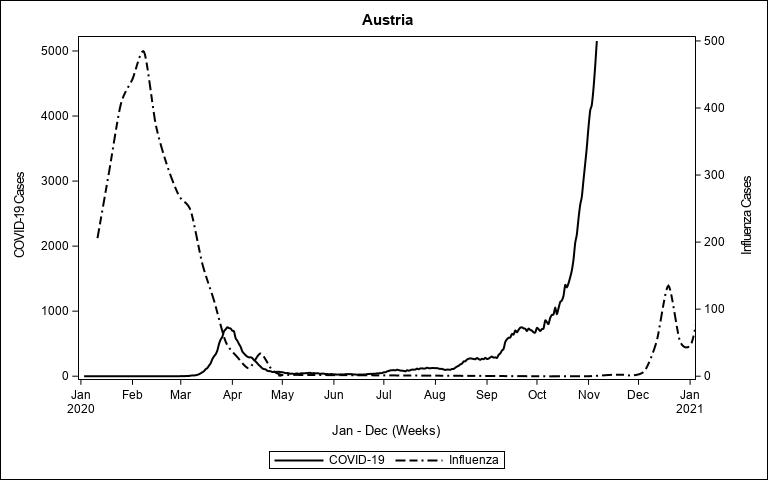

### Bahrain1.jpeg

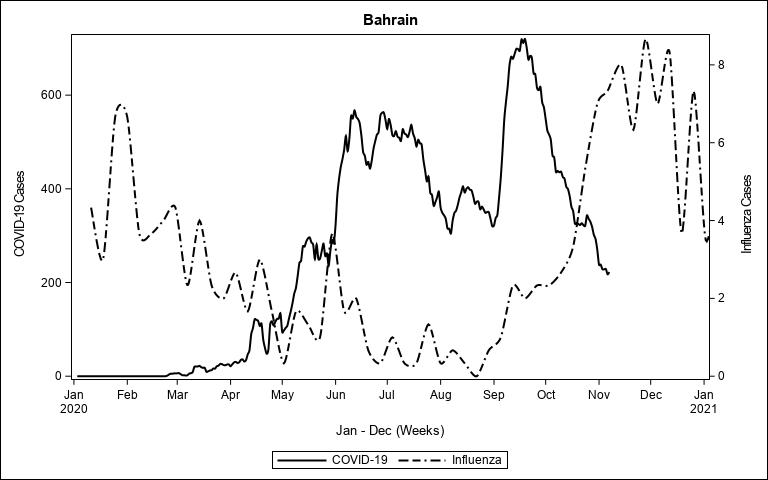

### Bangladesh1.jpeg

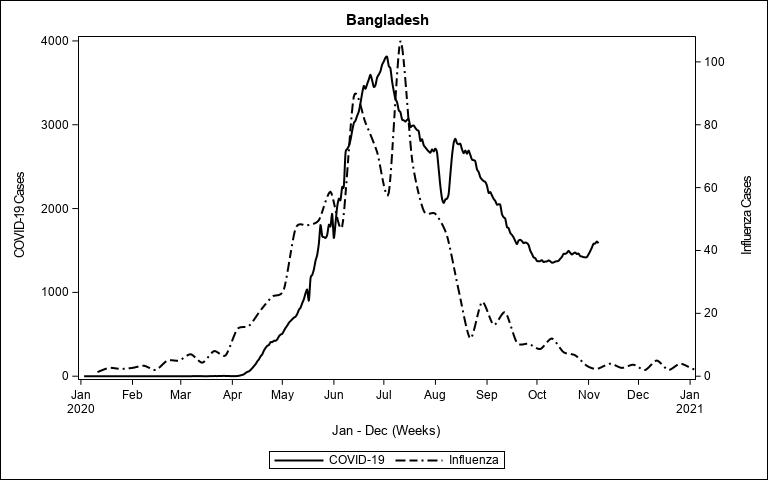

### Belgium1.jpeg

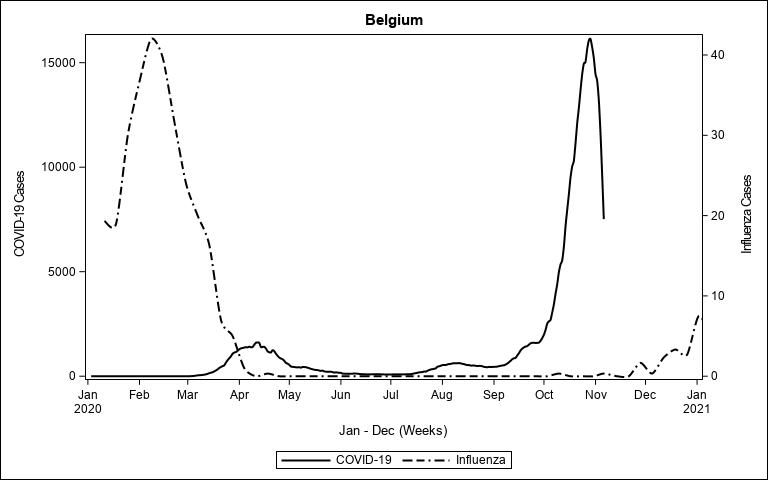

### Bhutan1.jpeg

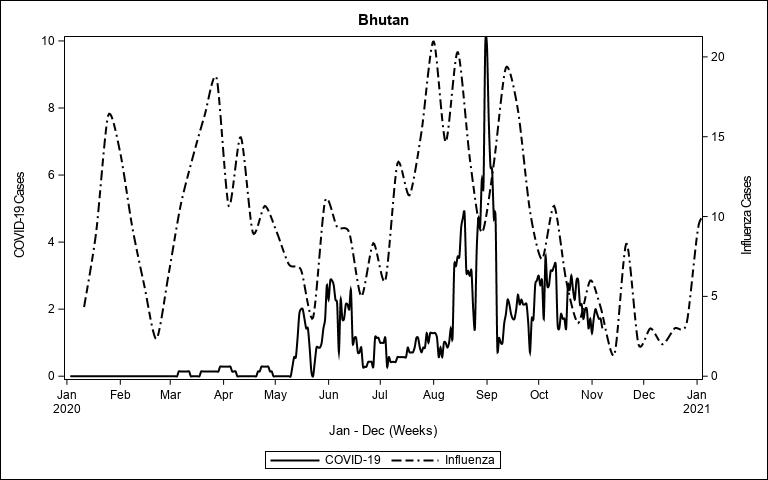

### Bolivia1.jpeg

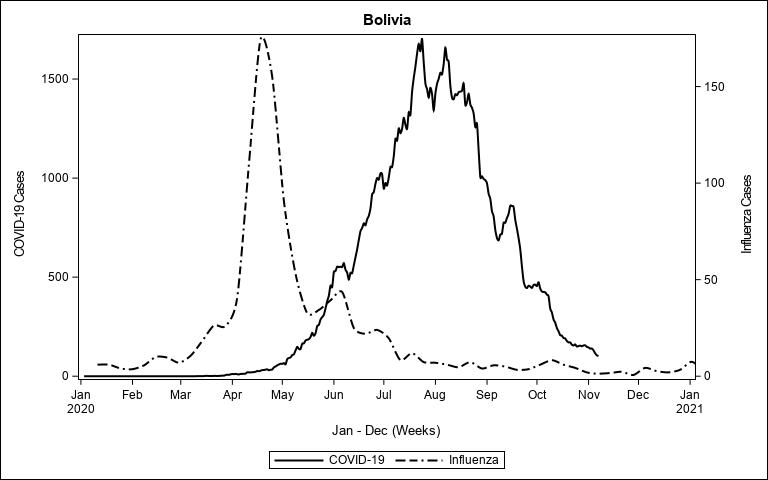

### Brazil3.jpeg

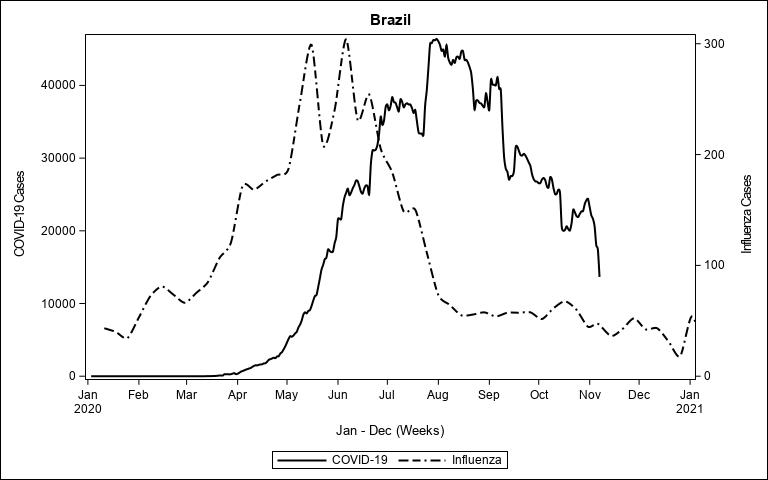

### Cabo Verde1.jpeg

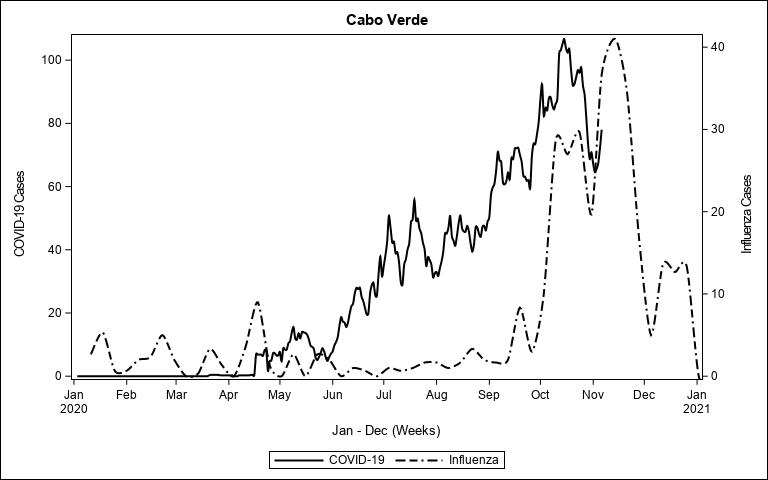

### Cambodia1.jpeg

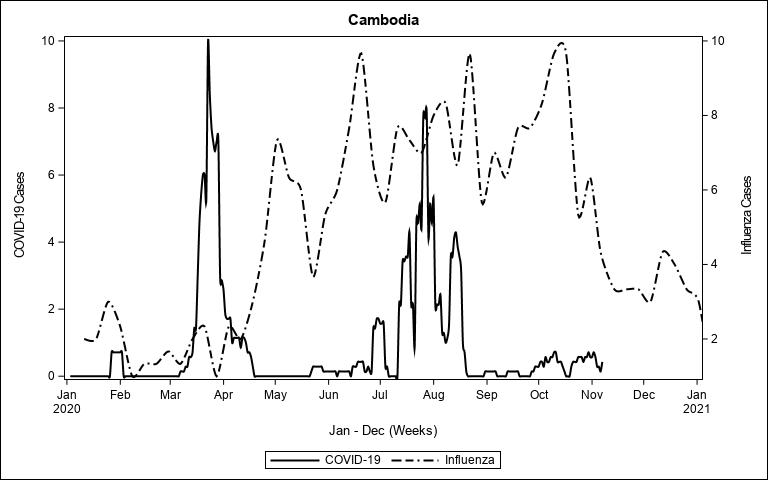

### Canada1.jpeg

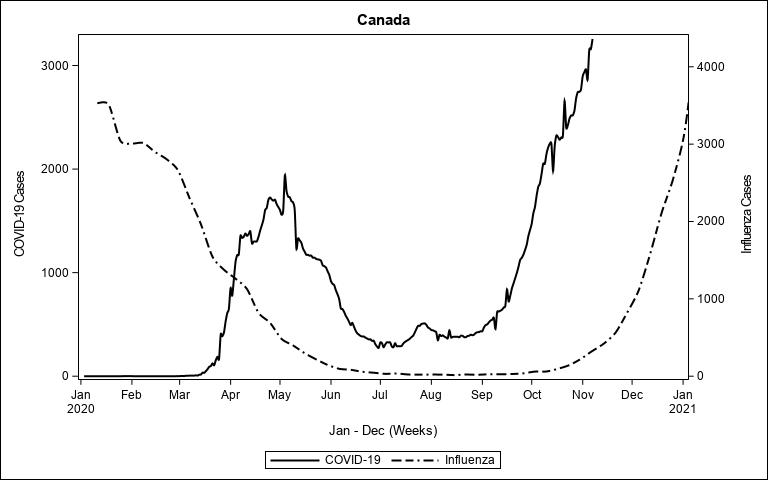

### Chile1.jpeg

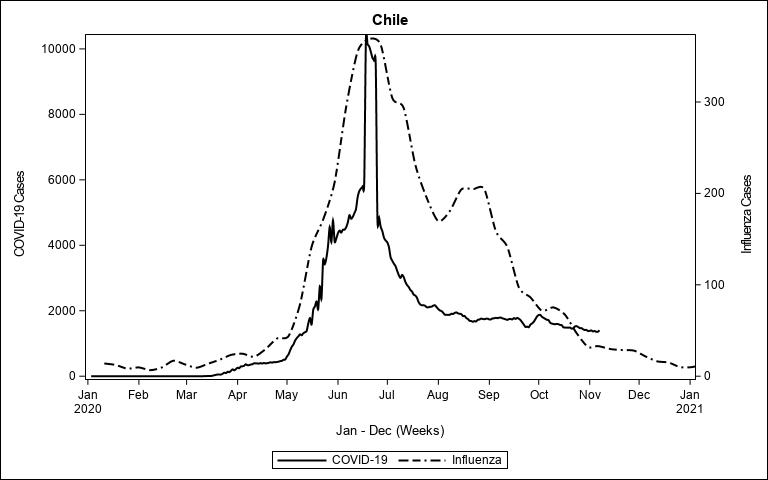

### China1.jpeg

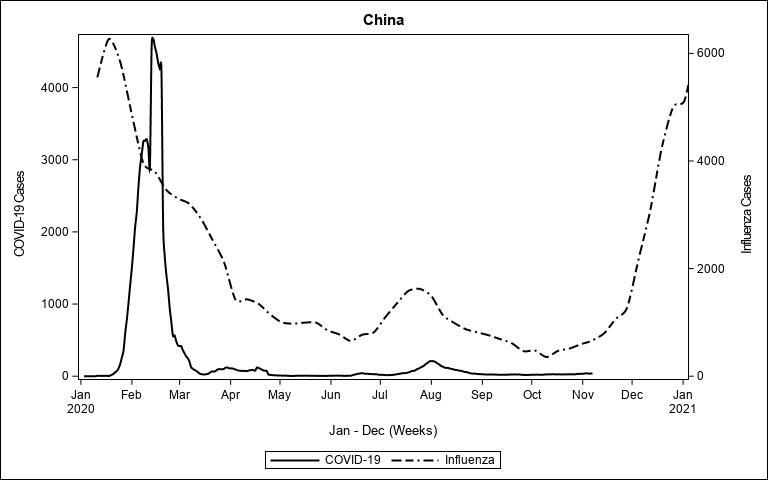

### Colombia1.jpeg

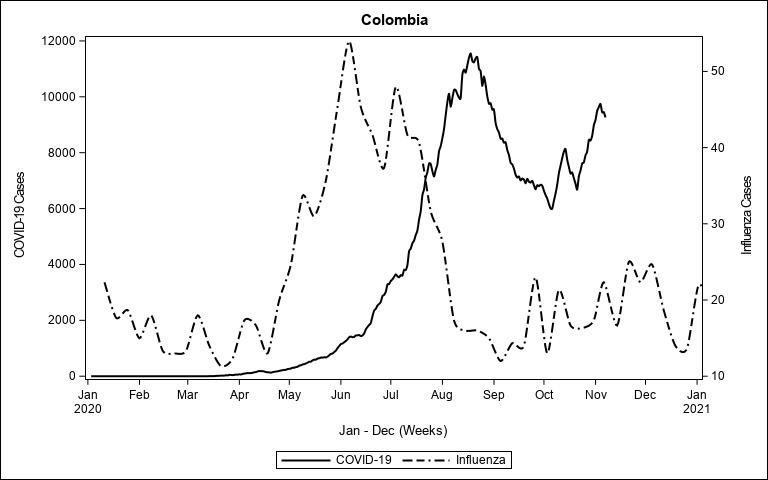

### Costa Rica1.jpeg

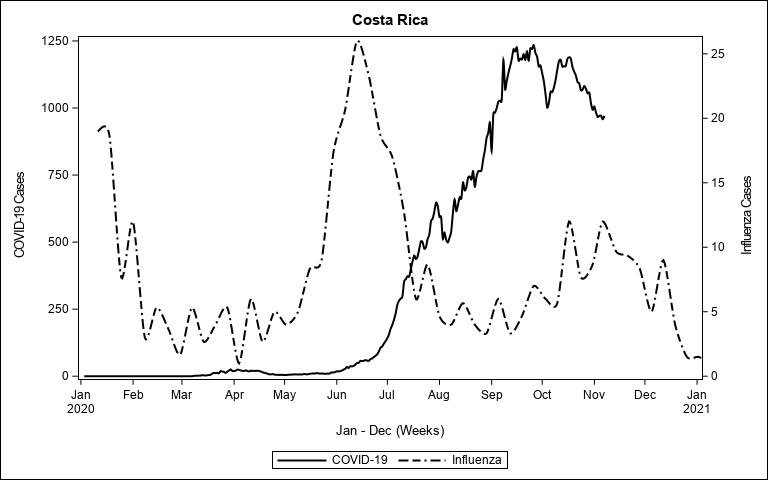

### Cuba1.jpeg

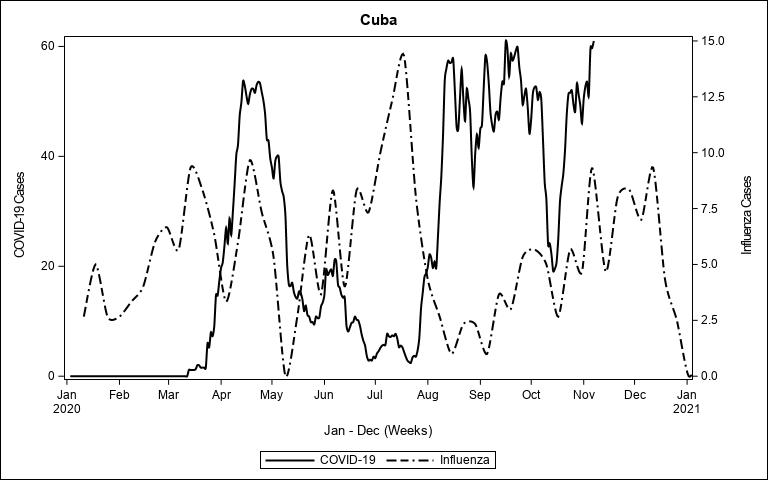

### Egypt1.jpeg

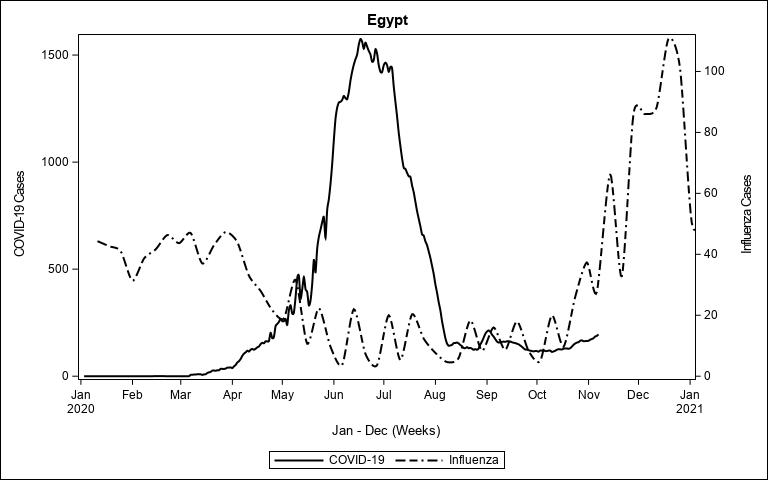

### El Salvador1.jpeg

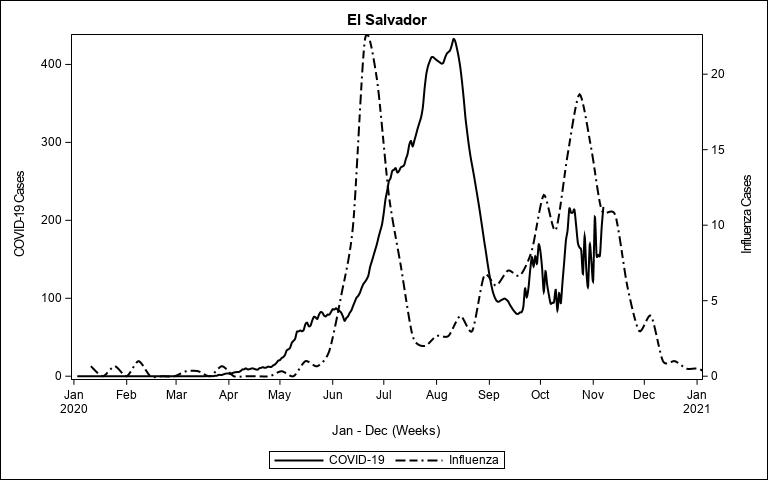

### Ethiopia1.jpeg

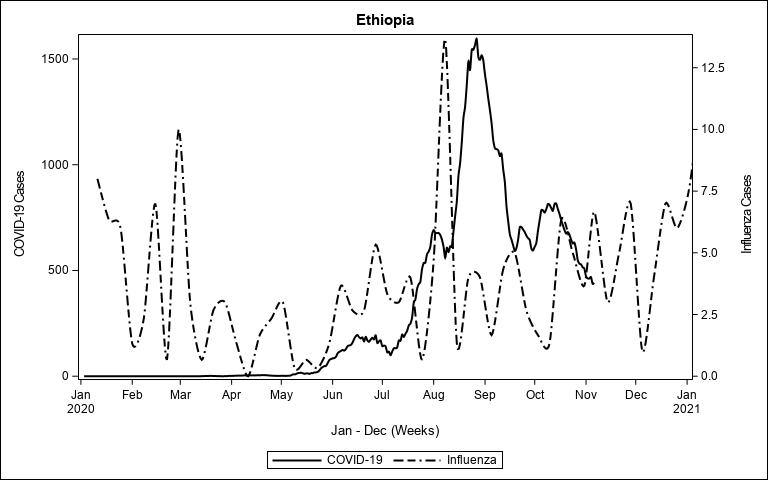

### Ghana1.jpeg

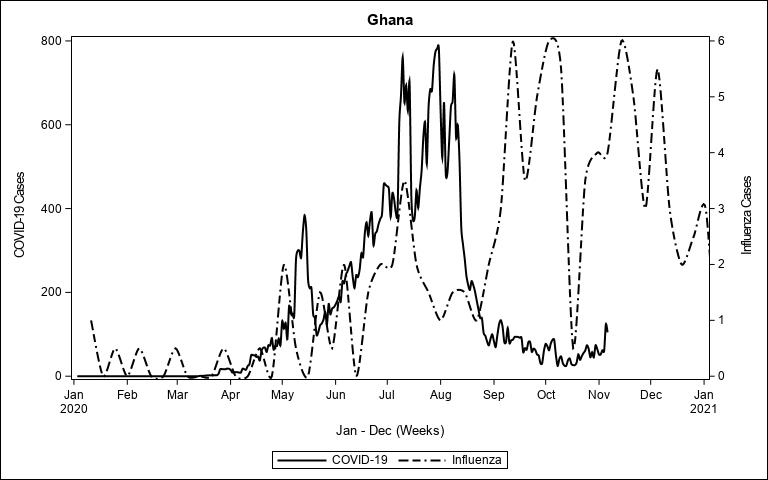

### Guatemala1.jpeg

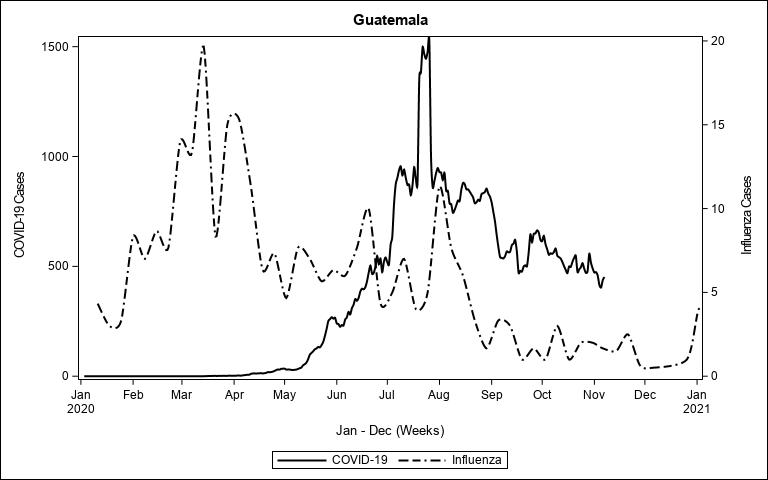

### Haiti1.jpeg

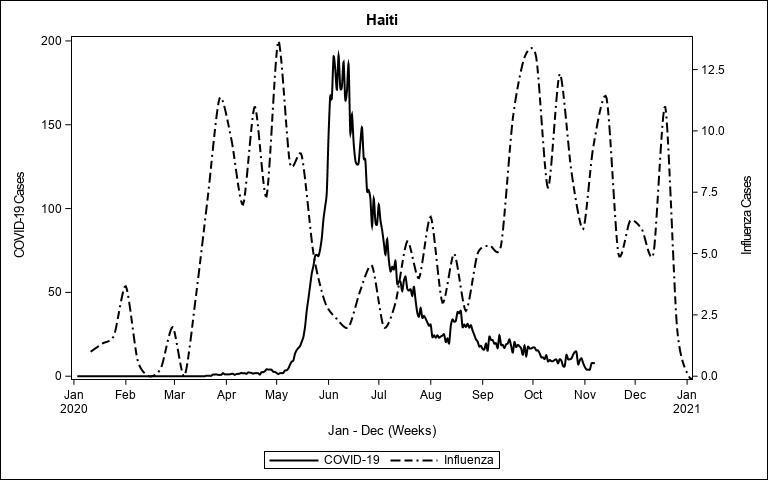

### Honduras1.jpeg

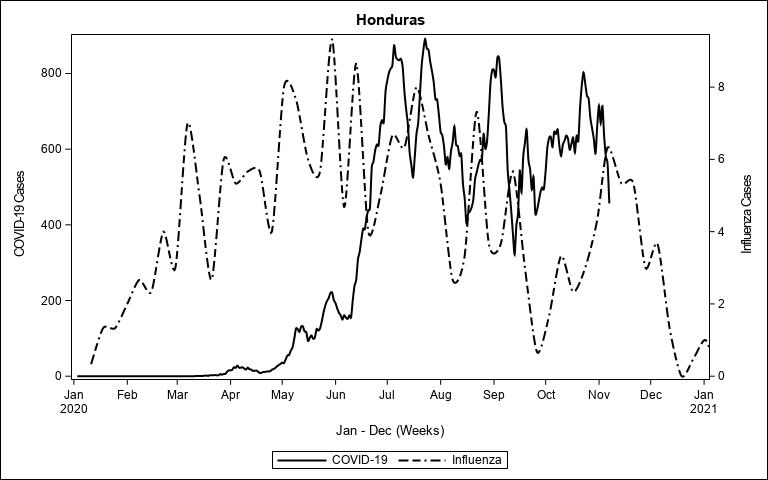

### India3.jpeg

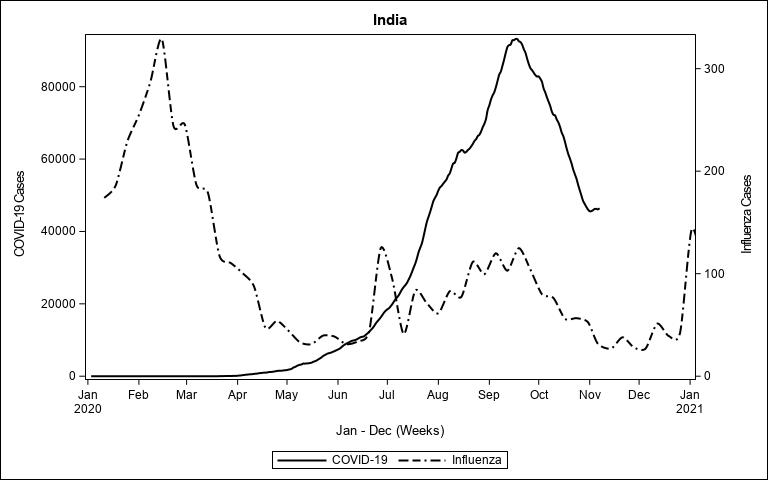

### Indonesia1.jpeg

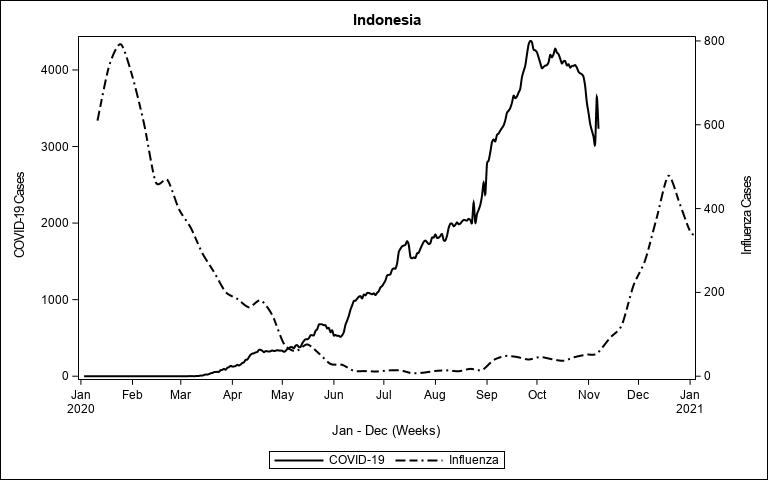

### Iran1.jpeg

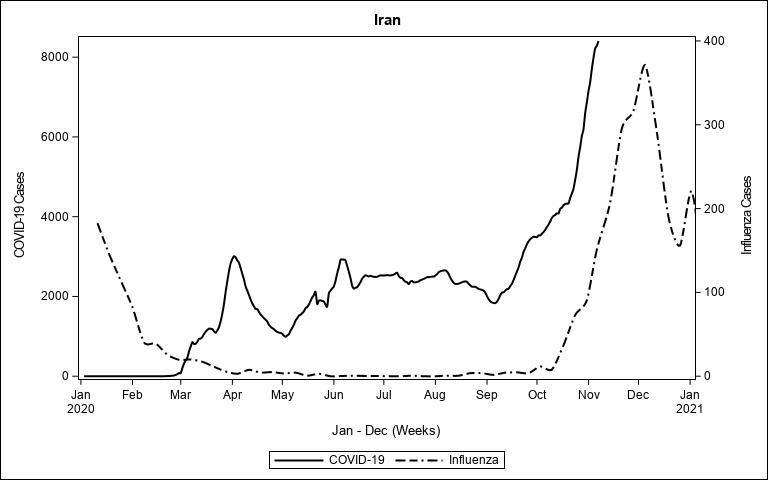

### Jamaica1.jpeg

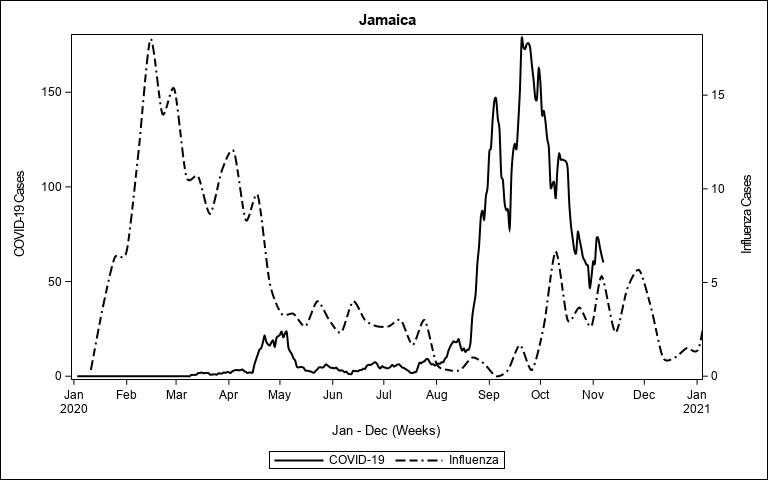

### Jordan1.jpeg

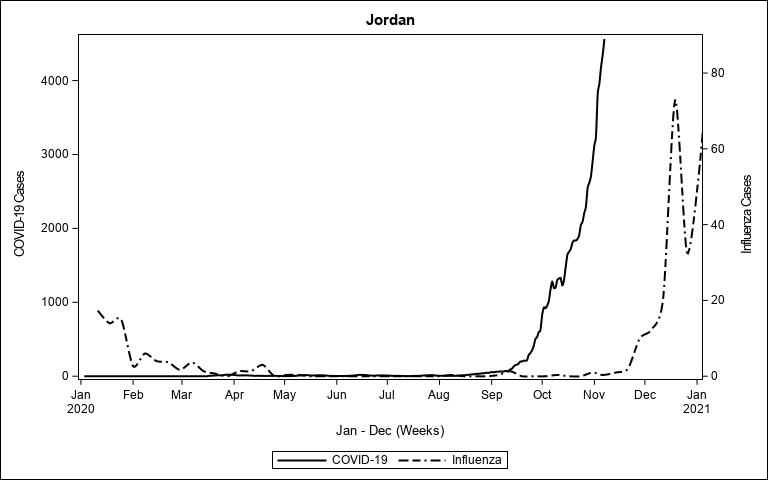

### Kenya1.jpeg

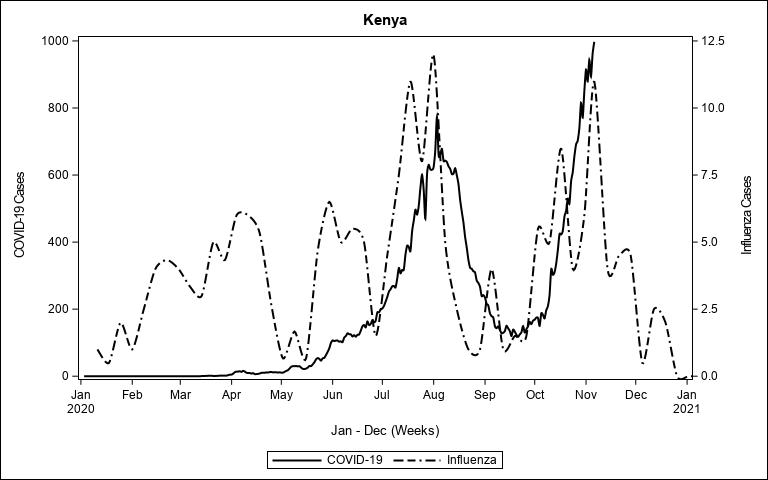

### Lebanon1.jpeg

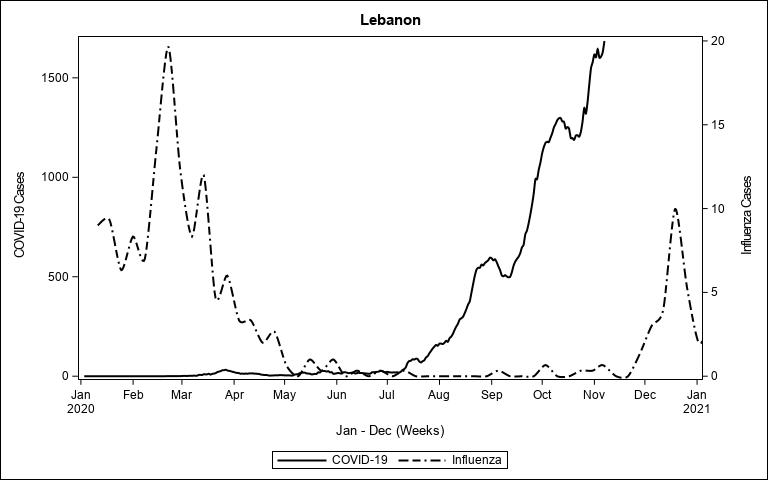

### Madagascar1.jpeg

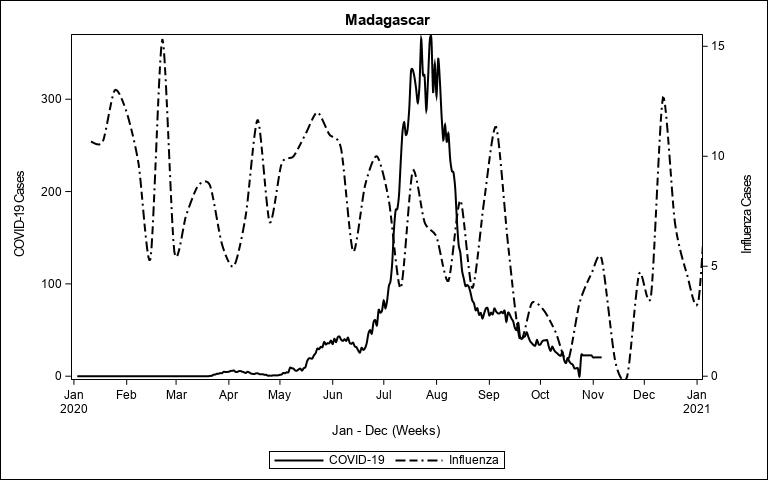

### Maldives1.jpeg

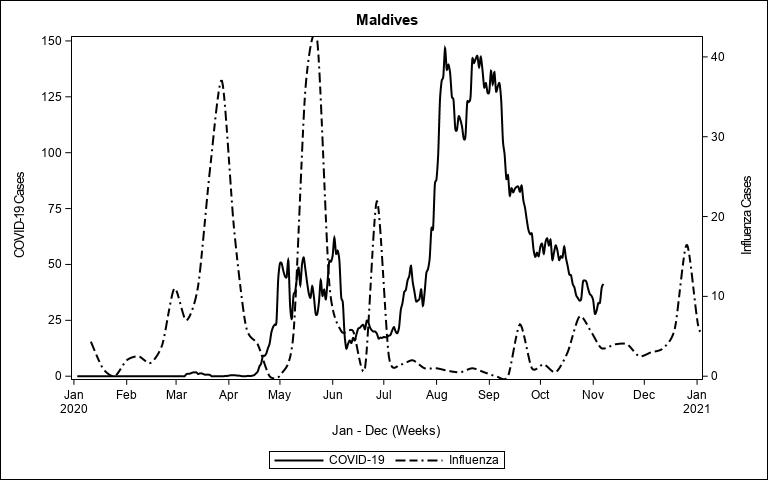

### Mexico1.jpeg

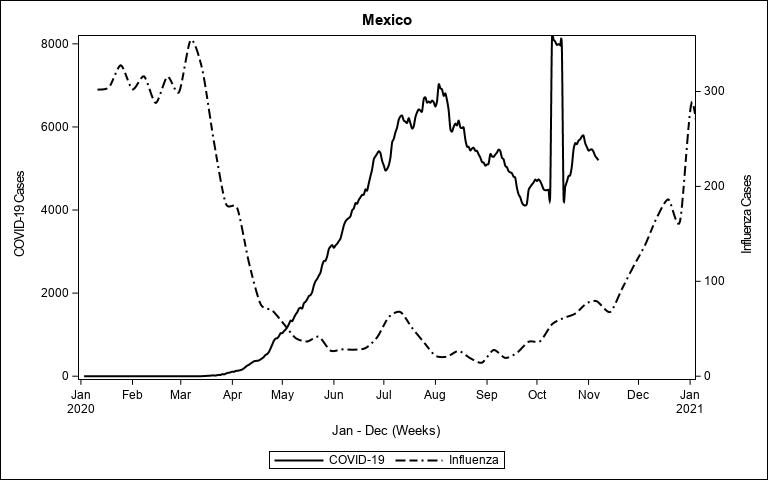

### Mozambique1.jpeg

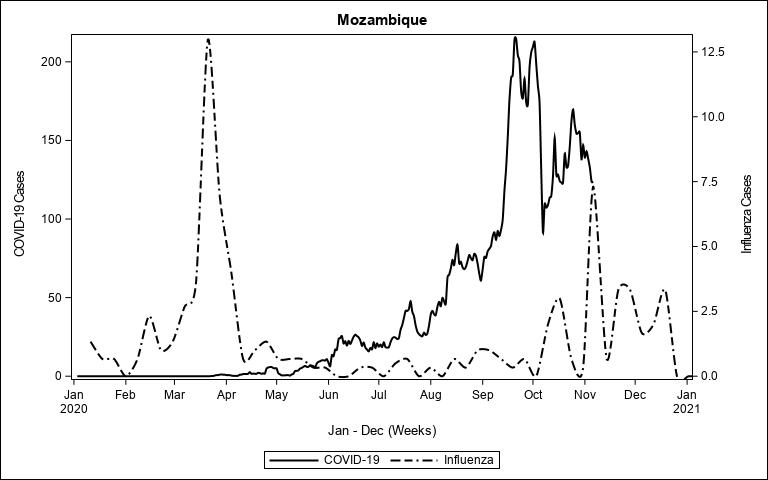

### Myanmar1.jpeg

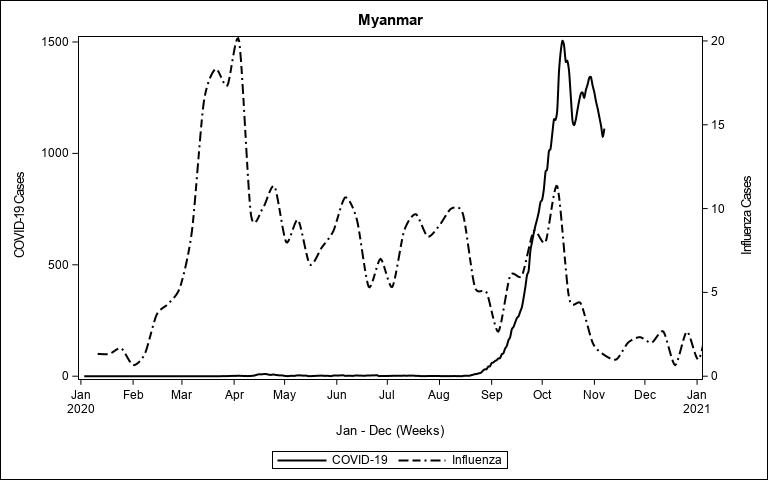

### Nepal1.jpeg

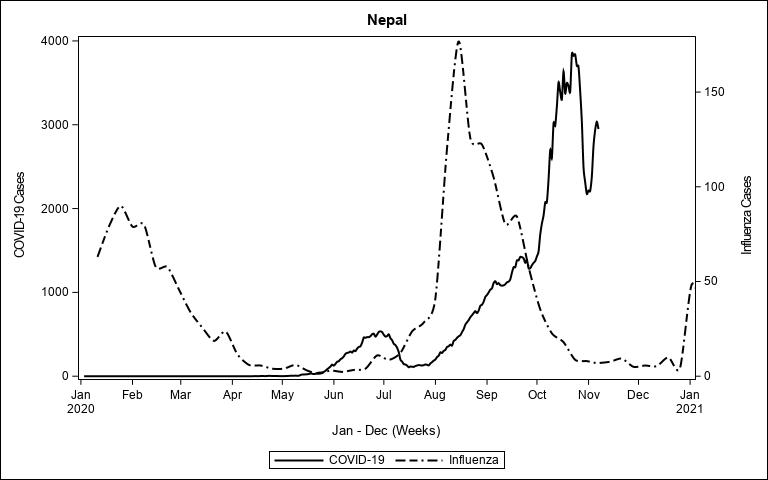

### Netherlands1.jpeg

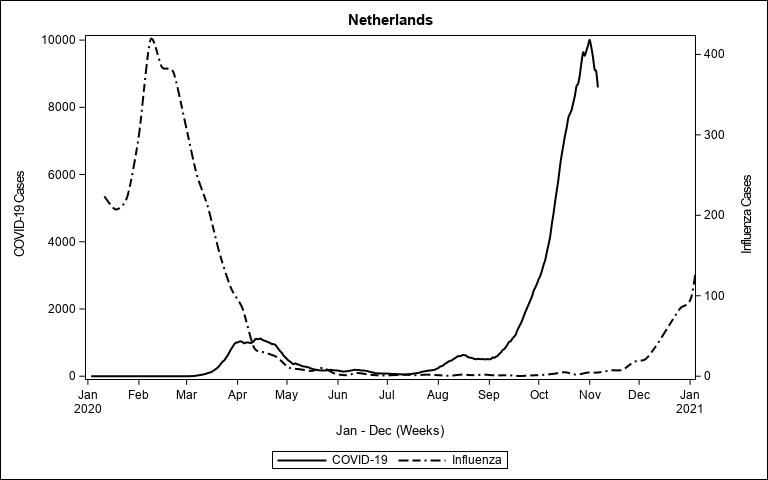

### Nigeria1.jpeg

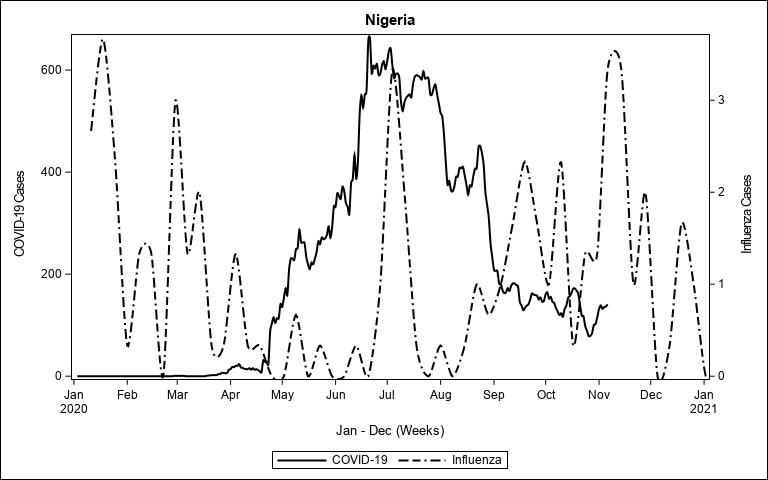

### Oman1.jpeg

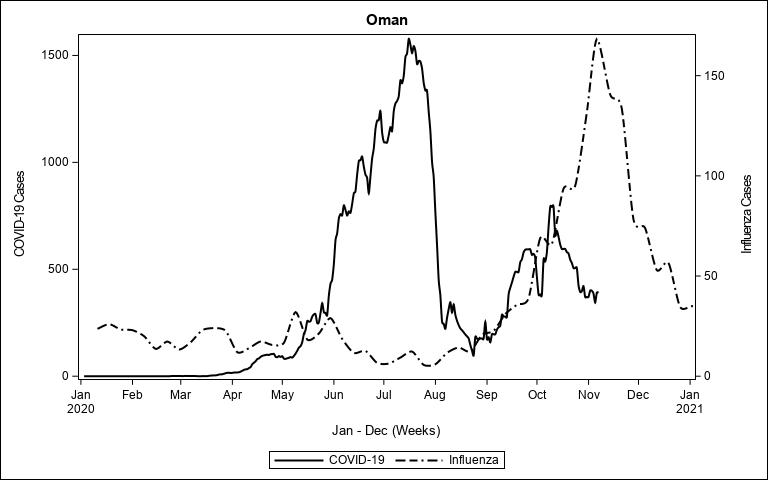

### Panama1.jpeg

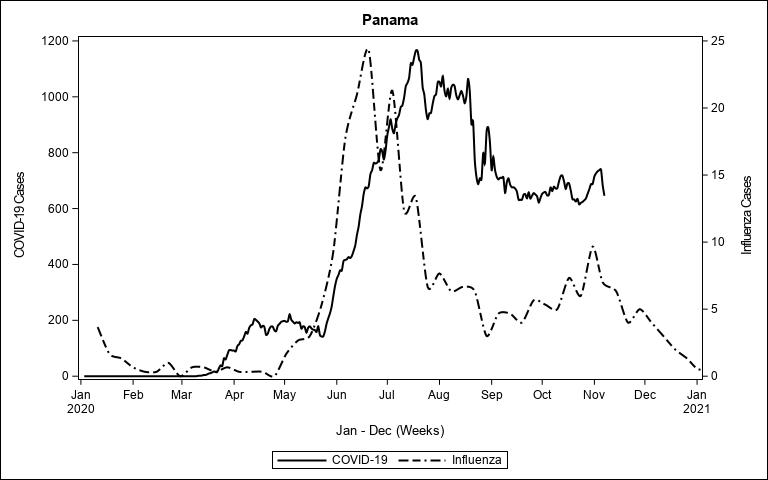

### Papua New Guinea1.jpeg

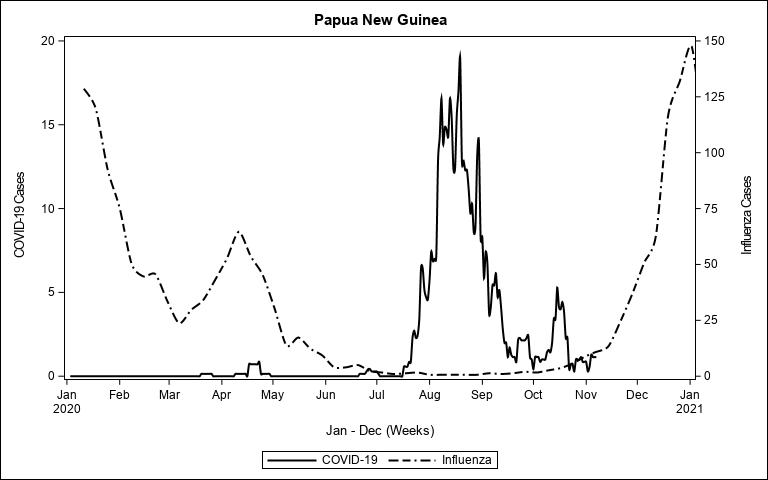

### Paraguay1.jpeg

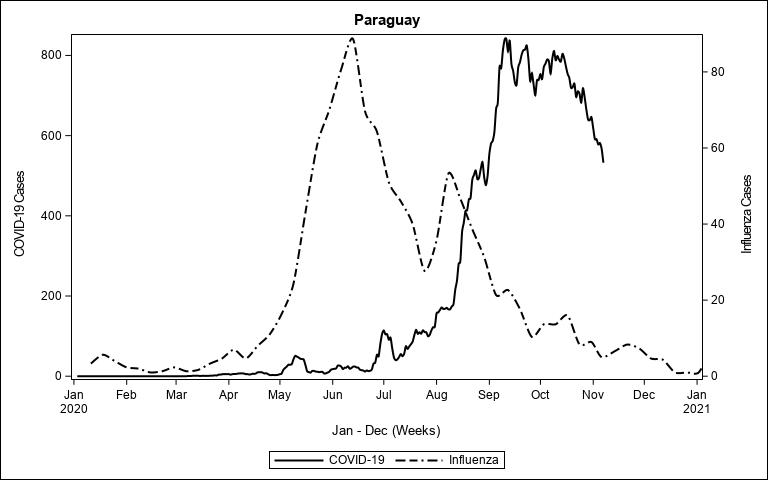

### Peru1.jpeg

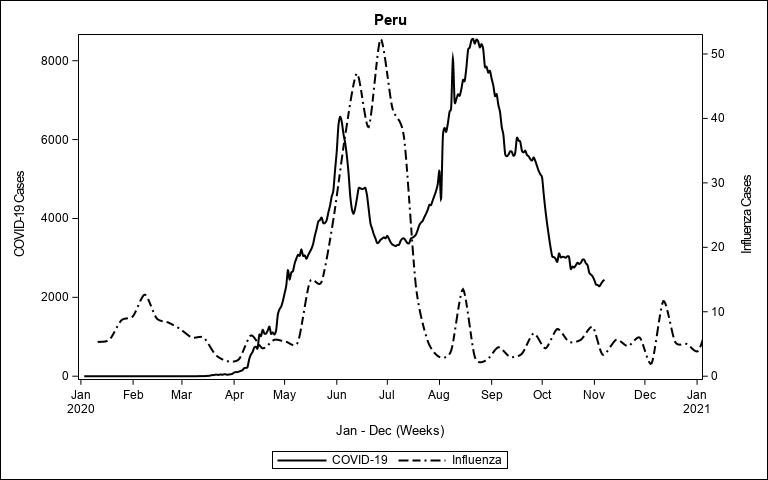

### Poland1.jpeg

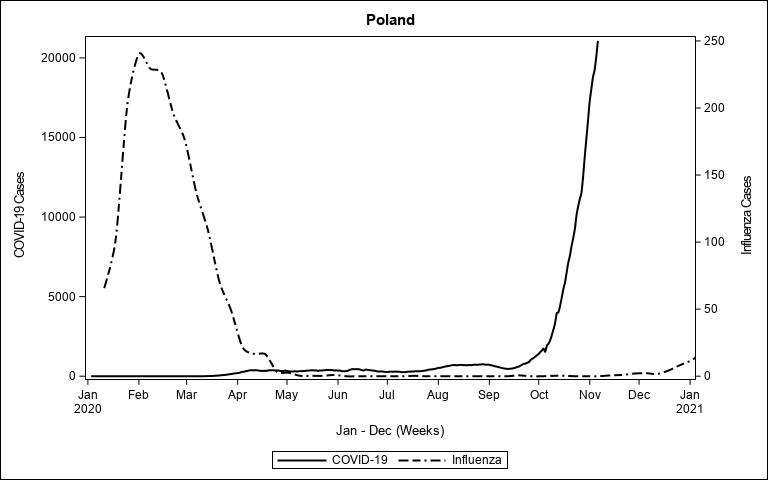

### Qatar1.jpeg

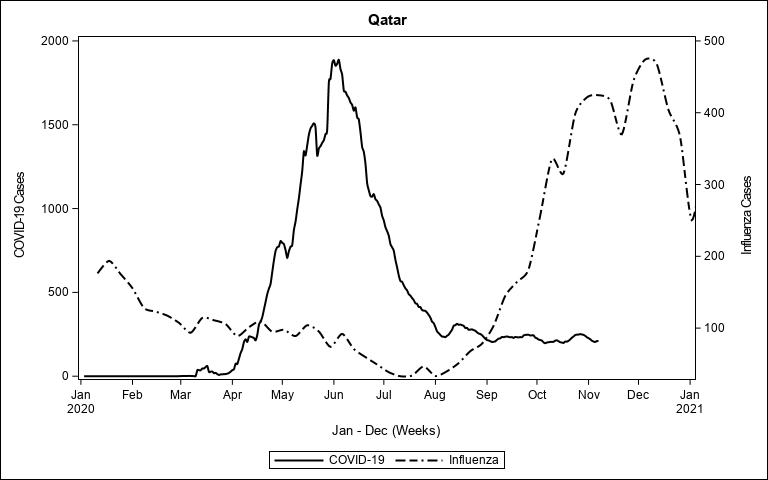

### Russian Federation1.jpeg

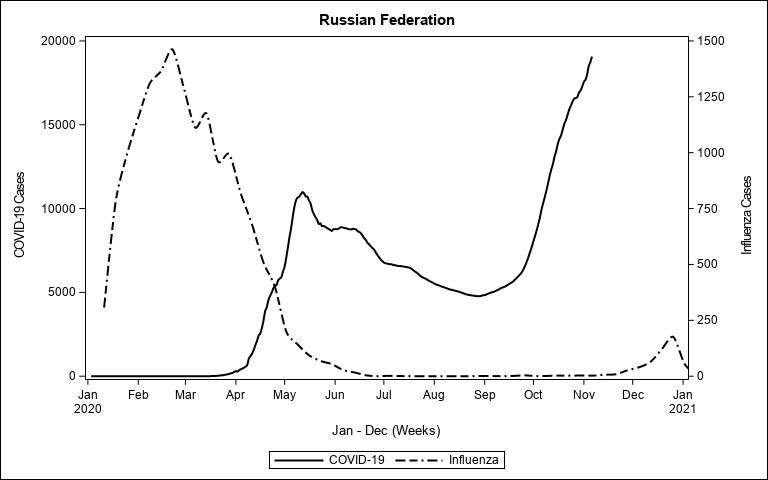

### Saudi Arabia1.jpeg

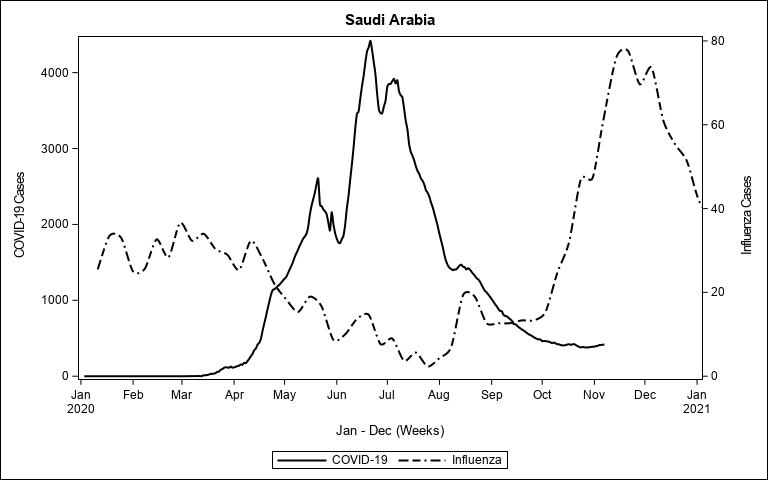

### Senegal1.jpeg

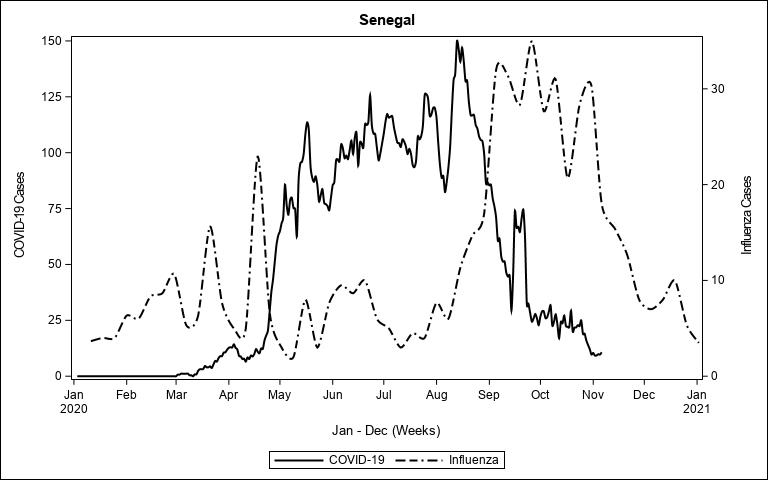

### South Africa3.jpeg

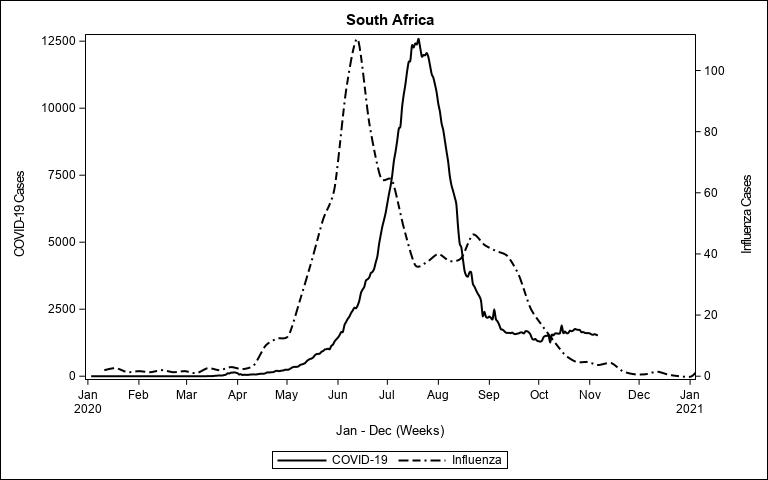

### Spain1.jpeg

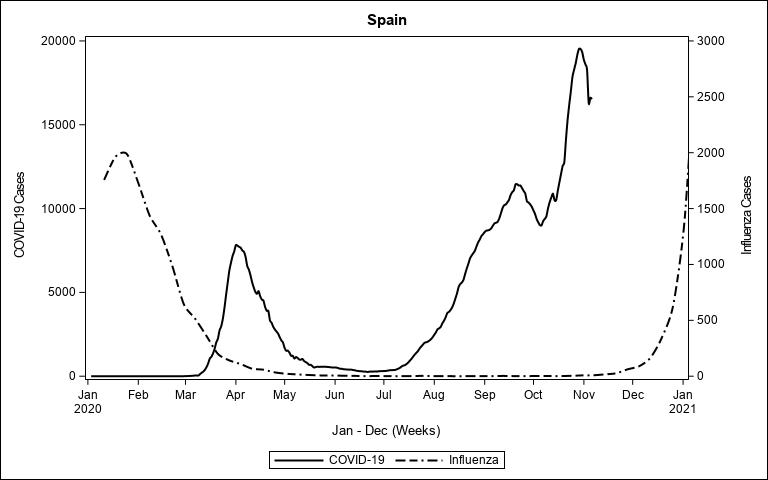

### Sri Lanka1.jpeg

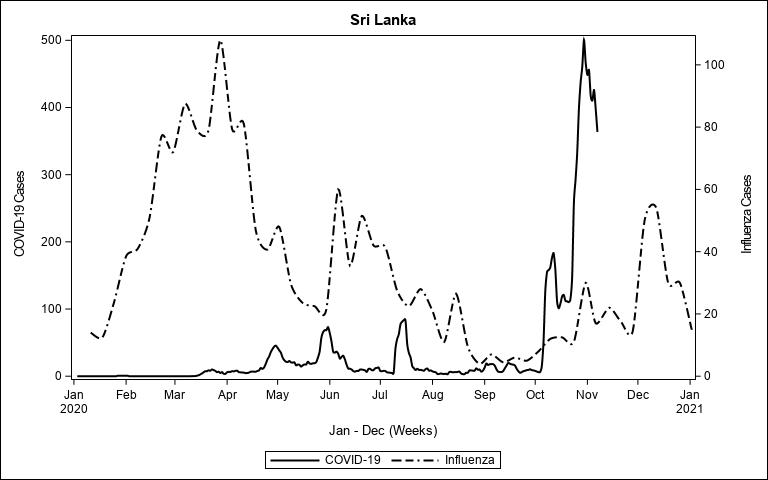

### Sweden3.jpeg

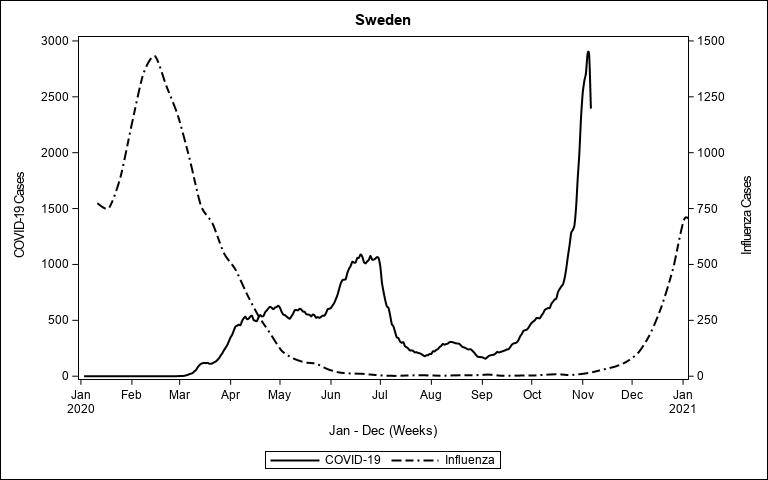

### Thailand1.jpeg

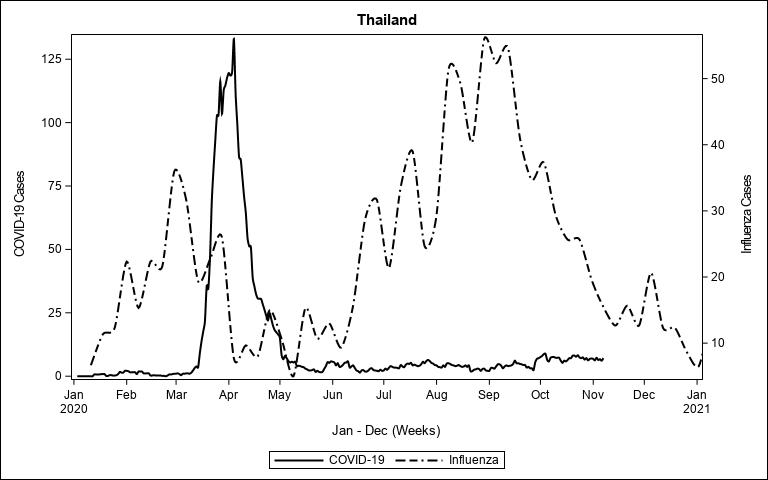

### Uganda1.jpeg

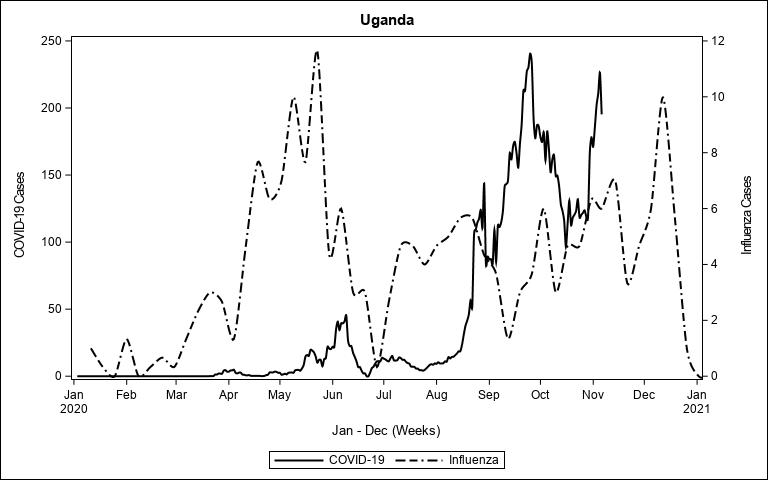

### United Kingdom1.jpeg

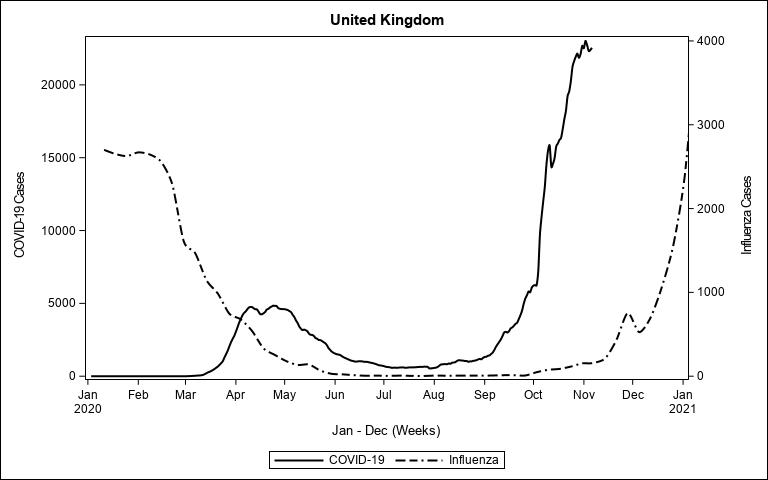

### United States of America3.jpeg

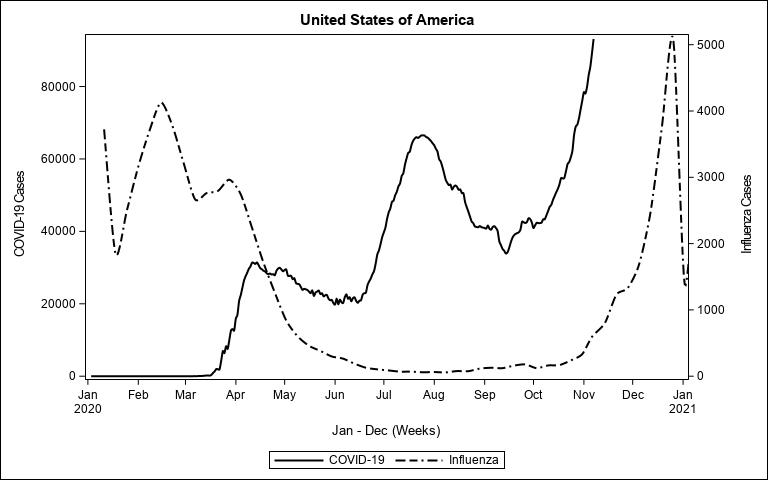

### Venezuela1.jpeg

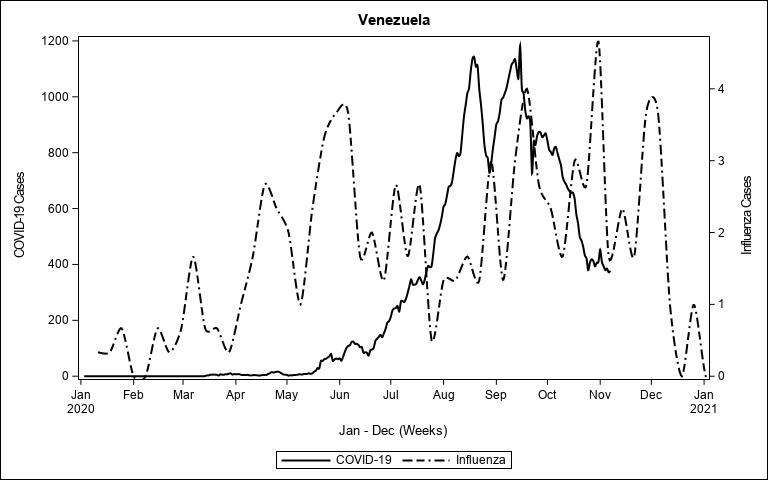

### Vietnam1.jpeg

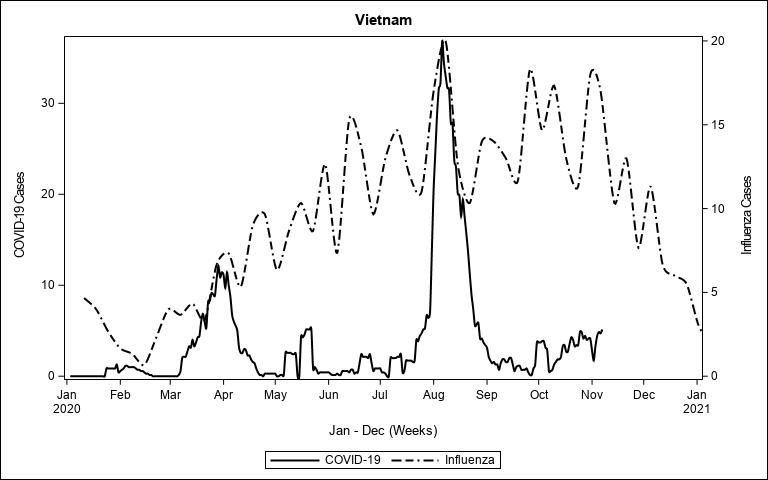

### Zambia1.jpeg

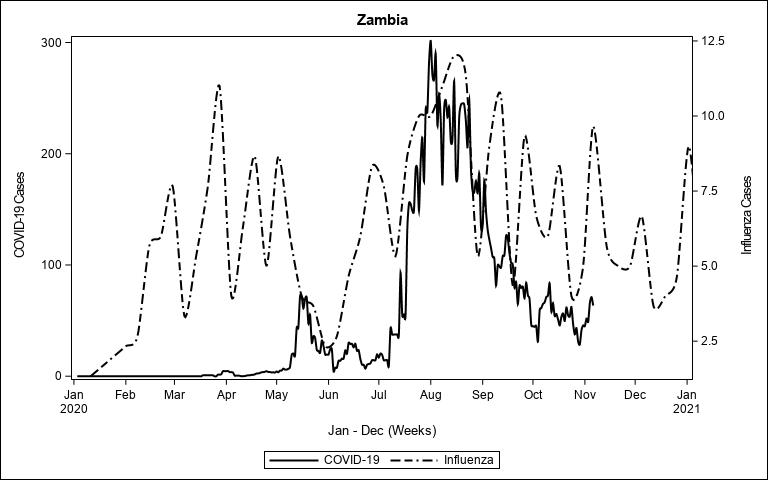
